# Applying a Socio-Ecological Model to Understand Factors Impacting Demand for Childhood Vaccinations in Nigeria, Uganda, and Guinea

**DOI:** 10.1101/2022.08.15.22278784

**Authors:** James Bell, Belinda Lartey, Gemma Spickernell, Natasha Darrell, Frances Salt, Cassie Gardner, Emily Richards, Lanre Fasakin, Shadrach Egbeniyi, Emmanuel Odongo, James Ssenkungu, Rigobert Kotchi Kouadio, Mamadi Cissé, Axel Bruno Ayiya Igowa Rérambyah, Maikol Adou, Rebecca West, Sunny Sharma

**Affiliations:** Ipsos Healthcare, 3 Thomas More Square, London E1W 1YW, UK; CMRG, 24A, Eric Moore Street, Wemabod Estate, Off Ajao road, Ikeja, Lagos, Nigeria; Ipsos Uganda, 3rd floor Padre Pio House, Plot 32, Lumumba Avenue P.O. Box 21571, Kampala, Uganda; Ciblage, Immeuble 5658, 2ème Etage, Rue 13, Liberté 5, Sicap Liberte, Dakar, Sénégal; Ciblage, En face de l’Ecole les Ecureuils – Lambanyi, Ratoma, Conakry, Guinée; Ciblage, Face SODECI, Imm. Hué, Porte B10, Riviera Attoban, Cocody, 09 BP 799, Abidjan 09, Côte d’Ivoire; Boston University School of Public Health, 715 Albany St, Boston, MA 02118, United States

## Abstract

Vaccines have reduced child mortality across the world, but low levels of demand for vaccination threatens to undermine progress. Existing frameworks to understand demand tend to prioritise caregivers’ decision-making processes. We aimed to build a wider understanding of vaccine demand by applying an adapted socio-ecological model to analyse 158 interviews with caregivers and fathers of young children, and community influencers in Nigeria, Uganda, and Guinea. We found that several factors come together to inform a caregiver’s demand for vaccination, including their familial and social relationships, their interactions with government and healthcare institutions, and the wider social and cultural norms in their communities. The study suggests that interventions targeted at families and communities instead of individuals could be effective. The results could be used to ensure that vaccine demand frameworks used by researchers and intervention designers are comprehensive and consider a wider range of influences on the caregiver.

## Introduction

Immunization is a highly effective public health intervention which has contributed to large global reductions in mortality and morbidity from preventable childhood diseases over the last half-century (Bloom, 2011). Significant gains have been made in extending the availability of vaccines in low-income countries, but there are many threats to achieving and maintaining high coverage, which risks resurgence of once-controlled diseases (Feikin et al., 2016).

Threats to vaccination coverage are often conceptualised either as problems with supply or demand, although there may be considerable overlap between the two (Muzumdar & Cline, 2009). Demand is typically defined as ‘the actions of individuals and communities to seek, support and/or advocate for vaccines and vaccination services’ (Hickler et al., 2017). Demand-side issues have received a lot of recent scholarly attention as international organisations such as the Gavi Alliance are increasingly focussed on this topic (*Annual Progress Report (2019)*, 2019).

Much scholarship on demand for vaccination in sub-Saharan Africa considers barriers caregivers face when seeking immunization services. This includes a lack of awareness and understanding of what vaccines do, distrust in vaccines and the broader healthcare system, other childcare or family priorities which may supersede vaccination, preference for religious or traditional modes of protection and the influence of family and community members which may inhibit vaccination-seeking (Cobos Muñoz et al., 2015; Dubé et al., 2013; Favin et al., 2012; Kestenbaum & Feemster, 2015; Lane et al., 2018; MacDonald, 2015; Majid & Ahmad, 2020; Mohanty et al., 2018; Yahya, 2007). Social and environmental factors are also commonly considered, including the influence of gender norms on health-seeking behaviour and practical difficulties in rural settings or conflict zones (Feletto et al., 2018; Lane et al., 2018; Levine et al., 2018; MacDonald, 2015; Okwo-Bele et al., 2018).

There are many examples of models which attempt to organise and consolidate the various elements affecting caregivers’ demand for vaccination. For example, the World Health Organisation (WHO) uses the COM-B model, which suggests that a caregiver must have the *capability, opportunity*, and *motivation* to vaccinate their child if their attempt is to be successful (Habersaat & Jackson, 2020). Models such as COM-B, while they do consider social and other contextual factors that may impact vaccination decision-making, tend to see vaccination as primarily an individual decision on the part of the mother. This may mean that models such as these do not engage with the full range of drivers and barriers, such as relationship dynamics, interactions with authority figures or social norms, that a mother may face.

Therefore, models examining demand for vaccination may underestimate the complexity of the issue by not incorporating a sufficient range of contextual factors, which may in turn impede the design of optimal solutions. This study aimed to describe the factors which create or undermine demand for vaccination among caregivers in more expansive terms, and in a way that explicitly engages with this topic as a systemic phenomenon.

## Methods

### Setting

The research took place in Nigeria, Uganda, and Guinea. These countries were chosen because of their differing vaccination coverage rates. Nigeria has achieved moderate coverage of the first and third DTP-HepB-Hib vaccines (DTP1 and DTP3), with 65.3% and 50.1% coverage respectively, but distribution is highly inequitable, with much lower coverage in the north of the country compared to the south (National Population Commission - NPC & ICF, 2019). Uganda has reached high coverage for DTP1 (94.9%) but has problems with ‘drop-outs’ for the later doses as only 78.6% have received DTP3 (Uganda Bureau of Statistics - UBOS & ICF, 2018). Guinea has the lowest coverage of DTP1 and DTP3 (62.3% and 40.2%), which may in part be attributed to disruptions caused by the 2014-2016 Ebola epidemic and the detrimental impact it had on the health system (Institut National de la Statistique & ICF, 2019; Suk et al., 2016).

### Data Collection

Interviews were conducted using semi-structured discussion guides (see Supplementary Materials), informed by a literature review, the results of a formative ethnographic study, and discussions with stakeholders in each of the three countries, including EPI representatives and government health authorities. Separate discussion guides were written for caregivers, fathers, and influencers (including grandmothers, religious leaders, and political/traditional leaders). Discussions with fathers and caregivers (lasting 90 minutes for caregivers and 75 minutes for fathers) covered their interpersonal influences, role as a caregiver to young children, and their beliefs and attitudes to traditional medicine and immunization, with emphasis on the reasons why they did or did not take their child for vaccination. Interviewers with influencers (lasting 45 minutes) covered their personal and familial situation, their role in the community and their beliefs and attitudes to traditional medicine and vaccination. The discussion guides were translated into Yoruba, Igbo, Pidgin and Hausa (Nigeria) and Luganda, Acholi and Runyankole (Uganda). In Guinea, the discussion guide was translated into French and the moderators interpreted the questions into Soussou, Peul or Malinké as required during fieldwork.

In Uganda and Guinea, the ethnographic study took place before the qualitative fieldwork, but in Nigeria they took place concurrently with some overlap in the samples between the two studies. Caregivers who took part in the qualitative phase only (n=36) were interviewed using the full discussion guide, and caregivers who also took part in the ethnographic research were interviewed using a shortened version (n=12). This article reports the results of the qualitative interviews only.

Interviews were conducted by trained moderators, who were briefed over the course of three or four days (in person in Nigeria, but remotely in Uganda and Guinea due to COVID-19 restrictions). Interviews in Nigeria and Guinea took place face-to-face but were conducted by telephone in Uganda (either during one extended session or three 30-minute sessions) because of COVID-19 restrictions.

Interviews took place in three regions per country, which were chosen in consultation with local stakeholders to ensure a spread of vaccination coverage rates, ethnic groups, and geographic areas. Participants were recruited by local researchers through convenience sampling in public places in each of the chosen regions. Primary caregivers (defined as those who have primary caring responsibility for the child, typically the mother) and fathers were eligible for the interviews if they were responsible for the care of a child between the ages of 2-4 (in Nigeria) or 1-3 (in Guinea and Uganda). The age criterion was changed after fieldwork in Nigeria to improve parental recall of vaccinations in the first year of life. If participants had more than one child in the target age group, the interview focussed on their youngest child in that group. Approximate quotas were set to ensure a balance on several factors, including immunization status of the child, income group, urban or rural setting and level of education (see Supplementary Materials for screening questionnaires). The definition of the influencer sample varied from country to country, and was determined after initial caregiver and father interviews, but comprised individuals who held a position of prominence in the community, such as religious, traditional, or political leaders.

Written informed consent was obtained from all participants. An honorarium for their time was provided. Uganda: Caregivers and fathers (USh 36,700/ USD 10), influencers (USh 5,500/ USD 1.50); Nigeria: All groups (NGN 4000/ USD 7.50); Guinea: All groups (GNF 369000/ USD 37). The research received ethical approval from Makerere University College of Health Sciences Review Board in Uganda (Ref: 724), the National Health Research Ethics Committee of Nigeria (Approval number: NHREC/01/01/2007-25/09/2019) and the Comité Nationale d’Ethique pour la Recherche en Santé in Guinea (Ref: 026/CNERS/20).

### Analysis

All interviews were audio recorded and transcribed into English. The analysis process centred on the caregiver transcripts, as we were primarily interested in influences on caregiver demand for vaccination. We used an adapted Socio-Ecological Model to code the caregiver transcripts and coded the father and influencer transcripts thematically to add contextual depth and detail.

The Socio-Ecological Model was chosen as an analytical framework because of its emphasis on the inter-relations between individuals, their environment and wider social context, which is pertinent to our research question (Bronfenbrenner, 1989; Jill F. Kilanowski PhD, 2017). Several different versions of the socio-ecological model exist in the literature (Golden & Earp, 2012; Newman & Newman, 2020; *The Social-Ecological Model*, 2022). We adapted these frameworks to ensure that the entire life context of our target respondents could be adequately encompassed.

Our model posits that decisions are affected by the interaction of four nested and interrelated systems. These are: the *microsystem* (the immediate environment in which the individual lives, including relationships with family and romantic partners); the *mesosystem* (the interrelationships between different microsystems, for example the individual’s spouse and their mother); the *exosystem* (the institutions, whether formal or informal, that the person interacts with); and the *macrosystem* (the social and cultural values and ideologies in a person’s context). There are two key differences in the SEM we adopted compared to versions found in the literature. We removed the personal system, as our research question focussed on contextual factors, not individual decision-making processes. We also had a different understanding of the exosystem, which is sometimes understood as the ways in which social settings interact indirectly with the person under consideration via their microsystems (Newman & Newman, 2020).

Transcripts were reviewed for completeness and quality and returned to fieldwork teams if needed to correct errors and omissions. They were then divided between five analysts (INITIALS REMOVED FOR BLIND PEER REVIEW) and coded in NVivo v.12. POSITIONALITY STATEMENT REMOVED FOR BLIND PEER REVIEW.

The caregiver transcripts were primarily coded to one of the four levels of the socio-ecological model. Under each domain, thematic sub-codes were created to capture the contents of each interview in more detail. A similar process was completed for the father and influencer interviews: the transcripts were primarily coded to capture drivers and barriers to vaccination, with thematic sub-codes under each theme. The coding team met regularly to align code-frames, resolve any areas of uncertainty, and to discuss the findings with the teams in Nigeria, Uganda, and Guinea.

The results are reported using the socio-ecological framework structure, with a narrative overview of the major themes which emerged under each domain and differences across countries noted.

## Results

### Sample Composition

A total of 158 interviews were completed. The interviews took place between October and December 2019 in Nigeria, between June and July 2020 in Uganda and between January and March 2021 in Guinea.

Roughly equal numbers of interviews were completed with caregivers and fathers of children who had received all vaccination doses, some doses, and no doses (Table 1). Fathers had a higher median age than caregivers in all three countries. More interviews took place in urban areas (reflecting the demographic composition of the target areas). Most fathers and caregivers had primary education or below in Guinea and Uganda, whereas the majority had secondary or tertiary education in Nigeria. Muslim participants made up a greater percentage of the sample in Guinea and Nigeria compared to Uganda. Influencer types varied by country, but religious and political leaders were interviewed in all three settings (Table 2).

**Table 1:**
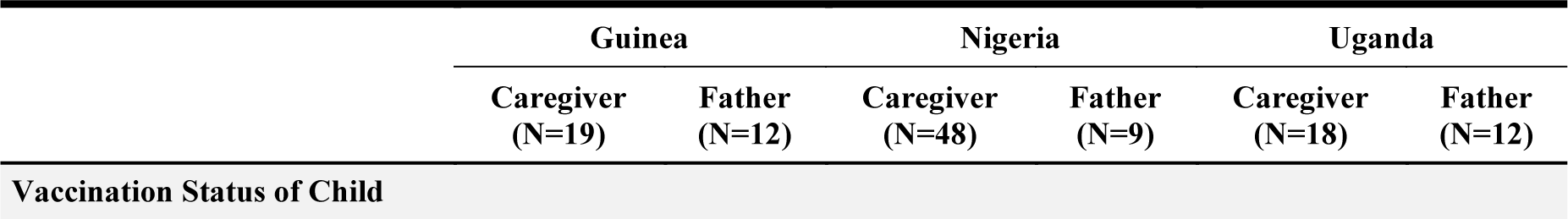
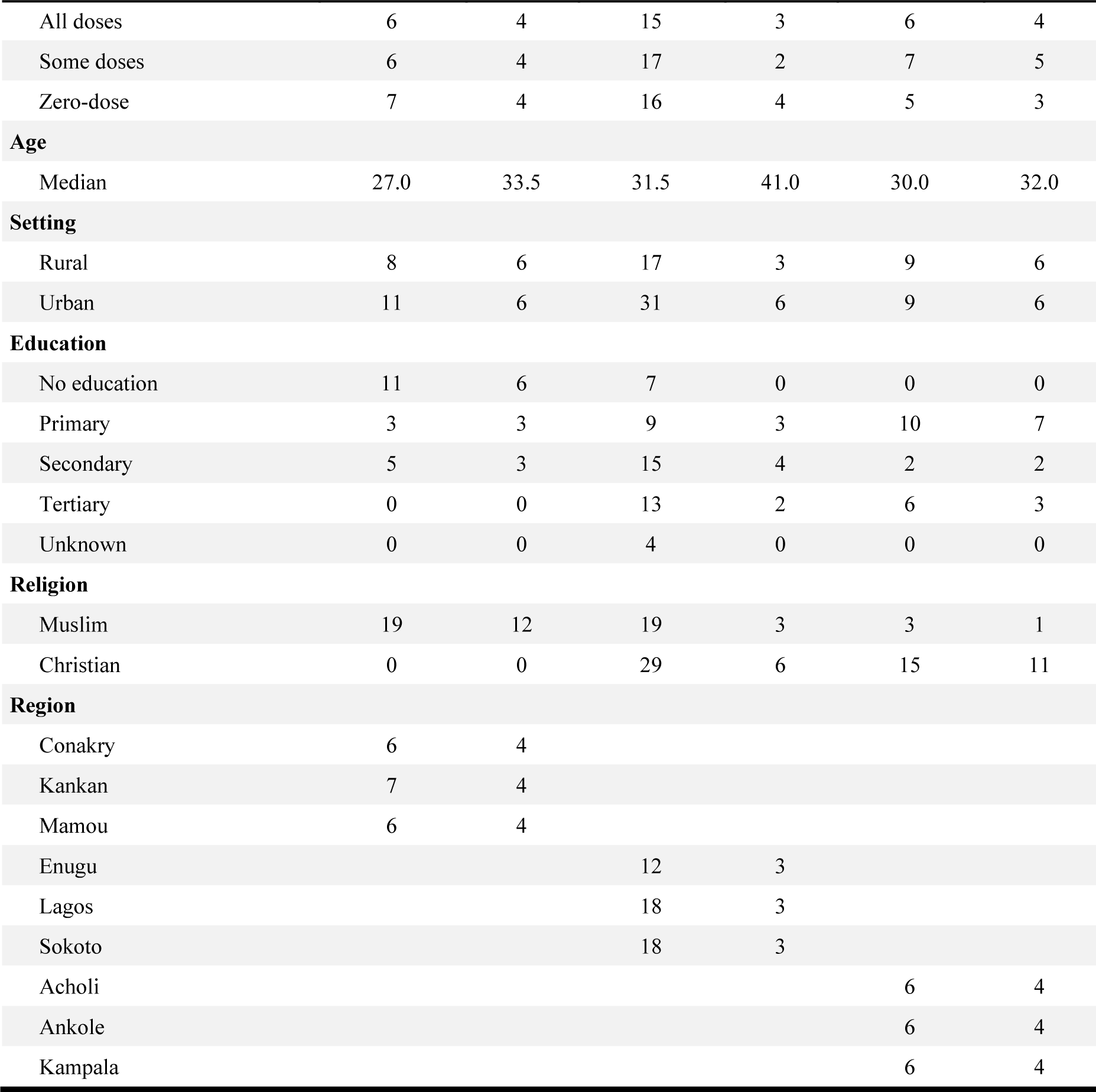
Description of Caregiver and Father Samples

**Table 2:** Description of Influencer Sample

|  | Guinea<br>(N=14) | Nigeria<br>(N=12) | Uganda<br>(N=14) |
| --- | --- | --- | --- |
| **Influencer Type** |  |  |  |
| Grandmother | 5 | 0 | 5 |
| Leader of women's association | 1 | 0 | 0 |
| Political | 3 | 1 | 4 |
| Radio presenter | 1 | 0 | 0 |
| Religious | 3 | 4 | 5 |
| Traditional | 1 | 7 | 0 |
| **Region** |  |  |  |
| Conakry | 5 |  |  |
| Kankan | 5 |  |  |
| Mamou | 4 |  |  |
| Enugu |  | 4 |  |
| Lagos |  | 4 |  |
| Sokoto |  | 4 |  |
| Acholi |  |  | 4 |
| Ankole |  |  | 5 |
| Kampala |  |  | 5 |

### Domains of the Socio-Ecological Model

A visual summary of the main findings is given in Figure 1.

**Figure 1:**
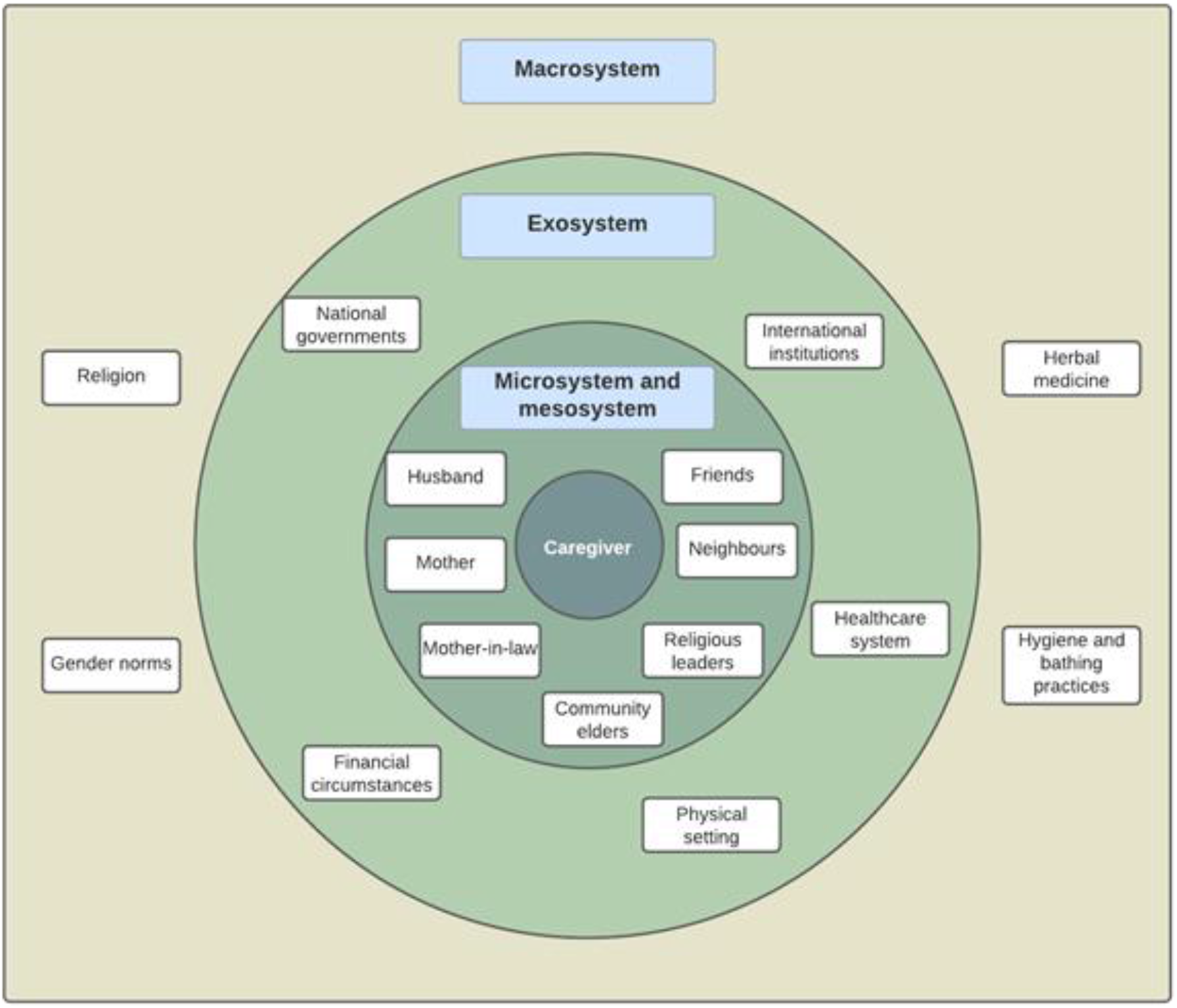
A summary of the main themes in the four levels of the model (2-column image, colour preferred)

#### Microsystem

The microsystem refers to the caregivers’ interpersonal relationships. Their microsystems tend to be large, encompassing immediate and extended family members, friends and neighbours, community elders or religious figures. Across study geographies, the most important individuals in terms of informing vaccination behaviour are the caregiver’s husband, mother/mother-in-law, community elders/religious leaders, and in some cases, neighbours, and friends.

### The caregiver’s husband

Husbands, as ‘heads of the household’, are near-universally recognised as the family’s primary decision maker. Wives can expect to be provided for materially by their husbands, but are expected to accept his decisions as final:

> *“[I respect my husband because] he is the crown over me. Like where I come from, respecting your husband means you have the fear of God and your parents in you […] The man that put you in [a] house, feeds you and does everything for you, after God and your parents, he’s the next”* (Caregiver, Nigeria)
>
> *“Knowing that the wife belongs to her husband, so she must do as her husband tells her to do”* (Caregiver, Guinea)

This extends to vaccination decision-making. Caregivers are expected to follow their husband’s wishes for vaccinating their children, and mothers have little recourse if they disagree:

> *“For me, I like immunization, but my husband is strongly against it […] I am under my husband, because he has even forbidden me to immunize his children”* (Caregiver, Nigeria)
>
> *“If [a wife] dares to defy the husband’s orders and go without his consent then she might lose her marriage”* (Caregiver, Uganda)

### The caregiver’s mother and mother-in-law

The caregiver’s mother (the child’s grandmother) is an important source of knowledge about child-rearing. In many ethnic groups included in the study (e.g., Igbo in Nigeria) it is commonplace for new mothers to be helped by their own mothers after the birth of a child:

> *“My mother was there for the omugwo* [an Igbo term for traditional postpartum care] *during which she bathes the baby, fed him and prepare meals for me and my baby. She also puts him to sleep when he cries and massaged my body and stomach with hot water and towel so that my stomach won’t remain big after healing”* (Caregiver, Nigeria)
>
> *“[My mother bathed the baby] since I couldn’t handle her because of the pain I still had. She cooked for us and took care of us in the first two months until I and the baby were strong enough to stay alone” (*Caregiver, Uganda)

Childrearing practices and traditions are passed on, which could include bathing techniques, the use of herbs to treat illness and advice on child nutrition. Immunization may be part of these discussions, which informs what action is taken over vaccination:

> *“I have known about vaccination since childhood because my mother sent me to get vaccinated”* (Caregiver, Guinea)

Caregivers’ mothers-in-law are also commonly present after the birth and also pass down wisdom and advice on child health, meaning they are also an important source of influence on vaccination:

> *“[I heard about vaccines] from my mother in-law. When I got pregnant she sat me down and advised me never to take her grandchildren for that vaccine because it caused a disability to one of her children. This boy had grown up crippled because of the polio vaccine. He received it at the age of 6 years and contracted polio which left him crippled, so I took her advice”* (Caregiver, Uganda)

The position of mothers in the communities often means that, as is the case with their husbands, caregivers are unable to go against their advice without suffering social or familial consequences.

> *“Everyone passes through me before doing anything. They ask my opinion on anything they want and I give my opinion along with prayers. When there is a problem in the household, I intervene by giving advice”* (Grandmother, Guinea)

### Community elders and religious leaders

Although their involvement and influence vary, community elders and religious leaders are often a source of advice for families with young children in practical terms:

> *“[People] should talk to an elderly person who is not a family member because that person can decide for both sides apart from the family member who may decide for only one side”* (Caregiver, Uganda)
>
> *“When there is a problem between two families, I intervene to solve the issue, I give them advice and show them what solidarity is, I tell them about my experiences in my home”* (Grandmother, Guinea)

This group may inform attitudes and behaviour around vaccination in two ways. Firstly, they can directly provide advice (or even directives) about vaccination, which carry weight because of their social standing:

> *“I’m the one who calls others to come with their child to take medication. When they come to vaccinate the children, I tell the women to take their children out to be vaccinated”* (Grandmother, Guinea)
>
> *“Even at church there is a pastor who visited, he said that only God can protect our children from diseases. Not even vaccines can, so the best thing to do for a child is to dedicate them to God because a lot of children have suffered due to vaccination”* (Caregiver, Uganda)

Secondarily, they may signal to caregivers and other family members what is acceptable behaviour in the community, or act as a model for community members to emulate. This can extend to vaccination:

> *“I am an elder in the community and the young people look at us as wise so we have a big task, we have to live an exemplary life in the community because younger mothers are there with all their eyes on us to learn from”* (Grandmother, Uganda)

### Neighbours and friends

These groups may also play a normative role in signalling to caregivers and other family members what is acceptable or unacceptable vaccination behaviour in the community. However, their chief role may be in bringing in alternative points of view, which caregivers can then build into their conception of vaccination. This may include stories of adverse events, vaccination rumours and conspiracy theories or other stories about the lived experience of vaccination:

> *“My neighbours say that vaccination is not good and my husband does not accept it, so we don’t agree to get vaccinated”* (Caregiver, Guinea)
>
> *“Once a while maybe when you see someone or a child that is disabled, the talk will start. Some will say it is because of […] vaccines, because they did not have it”* (Caregiver, Nigeria)

#### Mesosystem

The mesosystem describes how caregivers and fathers form opinions and make decisions (including about vaccinations) through the inter-relation of their various microsystems.

Many of the caregivers’ decisions take place at the interface of her relationship with her husband and her acquired membership of his family, and her relationships with her family of birth. As the quotes above demonstrate, through marriage she is considered to have become part of her husband’s family, but she also retains close links with her family of birth, which includes advice on child rearing.

One participant (Caregiver, Nigeria) described the tension between being responsible to one’s husband and to one’s birth family as *“an issue that disturbs every woman in this village”* when it comes to making decisions. There is evidence that the caregiver-husband microsystem takes precedence over the caregiver-birth family microsystem, including in relation to vaccination:

> *“I advised her not to go [for vaccination] so that it will not cause [a] problem in her home because there are some men, if they should give you instruction and you don’t obey them and if anything happens to her children there it may lead to divorce”* (Caregiver, Nigeria)
>
> *“It is better to lie to your mother than to lie or to disobey to your husband because we are in subjection to our husband and the manner in which treat our husband determines where we’ll spend our eternity. That is why anything that will make your husband get annoyed or upset don’t do it, you can know what to tell your mother but don’t disobey your husband*.*”* (Caregiver, Nigeria)

But these intersecting microsystems also provide opportunities for caregivers to covertly influence, or bypass altogether, decision-making processes that are dominated by their husbands. One respondent (Caregiver, Nigeria) recounted that her husband had forbidden her from taking her children for vaccination. Knowing that her mother-in-law was supportive of vaccination, she enlisted her support to covertly arrange for them to visit a clinic for immunizations. In this way, the interaction between microsystems produces conflict, but a caregiver may make use of her membership of overlapping microsystems to take some level of decision-making control on the topic. These negotiations often involve the caregiver’s parents or parents-in-law, but can also include other community members who can be called upon to exert their influence. In response to a question about how to resolve a hypothetical marital dispute, a caregiver in Uganda described how she would make use of her husband’s respect for community elders to influence him:

> *“Definitely, she needs to involve someone because they have failed to agree; the situation is above them. They need an older person to counsel them […] I trust my pastor to handle such issues [*…*] they need an older person like an auntie or uncle. Those have seen it all and the husband will respect them more than the siblings who could be younger or age mates and might despise them*.*”* (Caregiver, Uganda)

#### Exosystem

The exosystem includes formal and informal social and economic structures which have an impact on attitudes and behaviours.

Confidence in national governments and international institutions is low across the three countries, particularly in Guinea where it appears that the response to Ebola further eroded trust in public institutions. Vaccines can be understood by caregivers as extensions of the perceived inefficacy of the state to provide for the population, or as symbolic of encroachment of international institutions on domestic affairs. For some families, this can reduce trust in vaccines specifically, and the healthcare system more generally to distribute them:

> *“When Ebola started in 2014, I was in Siguiri, there was a person there who had nothing, his mother had come from Conakry, he sent him to the hospital, the doctors said that his mother was a suspect case, then they told the man that he must also be vaccinated if not he will contract the disease, the man was vaccinated, when he came back home a few days later, he was feeling dizzy, as soon as he was sent to the hospital they said he was contaminated, whereas a few days before he was healthy, since then I have withdrawn from vaccination”* (Father, Guinea)
>
> *“Because we can’t afford hospital bill, so why is government always insisting on immunization? See now, we are very poor, see where we are staying and government is not doing anything to help us. When you’re sick, government won’t provide any help but when you’re healthy, then government wants to help you with free immunization”* (Caregiver, Nigeria)

The experience of healthcare systems also reduces demand for vaccination. Inaccessible facilities, long queues, frequent stock-outs, poor experience of interactions with healthcare workers, and lack of information about side effects of vaccination, reduce caregivers’ desire to return for future doses.

> *“I was so disappointed when the health worker asked me to pay for the vaccine and yet I knew it was for free. I would rather go to a private hospital where the health workers are willing to give me attention, sit with them and get all these details because in these government health centres these health workers work as if they are rushing for something. They do not have time to explain everything to mothers”* (Caregiver, Uganda)
>
> *“How they attend to people or the way all those nurses talk at times can discourage people”* (Caregiver, Nigeria)

The caregiver’s physical and financial circumstances, which may be considered part of the exosystem, also have an impact on demand for vaccination. Caregivers across all three countries report that vaccination sites are sometimes unsafe to access due to poor transport infrastructure or high levels of crime, or too expensive to attend regularly due to transport costs or the need to pay healthcare workers for services. Immunization is considered by many to be a low priority in comparison to the caregiver’s need to earn money and care for their families, particularly in Uganda, where caregivers are more likely to be single parents.

> *“The biggest challenge is the distance, our health centre is very far and transport means are very scarce and costly and walking is so difficult so most parents try but fail along the way meaning they don’t complete all their vaccinations”* (Caregiver, Uganda)
>
> *“Some people are usually busy; probably they are so caught up with work and because they don’t have a maid, they fail to make it for the appointment on the immunization card”* (Caregiver, Uganda)

#### Macrosystem

The macrosystem encompasses the cultural values, traditions and norms which influence a parent’s demand for vaccination, including religion, traditional medicine, and hygiene.

### Religion

In each of the three countries, some parents say that they refuse vaccination because they believe that protection from God is enough to ensure that their child does not get ill. This sentiment was particularly strong in Uganda.

> *“I got fed up of all that stress [going for vaccination], and I decided to trust the Lord to protect my children when I got saved and learnt the fact that Christ heals, protects and holds our life in His hands regardless of what we do in our human nature”* (Caregiver, Uganda)

Religion impacts demand for vaccination in other more indirect ways. In all three settings, religion can reinforce other social norms, which in turn have an impact on vaccination behaviour. For example, many report that religious teachings reinforce the authority of a husband over his wife, which then makes it more difficult for a female caregiver to contradict her husband on vaccination matters. In this way, religion becomes important not for its direct impact on vaccination, but on how it influences relations at the levels of the microsystem.

> *“Islam teaches that you shouldn’t do what your husband has forbidden you from doing”* (Caregiver, Uganda)

### Traditional practices

In some settings caregivers use traditional practices to protect their children from disease. However, as religion can function to reduce vaccination demand in more circuitous ways, belief in traditional protection methods can feed into social systems which interact with vaccination.

Herbal medicine use is viewed as a pillar of tradition and the authority of elders. Seeking vaccination services can be interpreted as a rejection of traditional forms of knowledge and authority, which are typically perpetuated by community elders. To avoid this conflict vaccinations are either avoided or seen as interchangeable with traditional forms of protection.

> *“I go and look for herbs because they work. We have been taking herbal medicine since we were kids”* (Caregiver, Uganda)

Hygiene and bathing practices are commonly used as a form of child protection (particularly in Guinea). Through association with Islamic rites, these practices have gained cultural and religious legitimacy and thus may take precedence over newer protection methods (e.g., vaccination) that are perceived as ‘Western’ or external to the established community value system.

> *“Moderator: Are there ways to protect your child from these diseases?*
>
> *Respondent: What I know is cleanliness, to protect a person from illness, you have to be clean, when you make the child clean and watch what he eats, he will be more resistant to illness*.*”* (Caregiver, Guinea)

## Discussion

This study used an adapted version of the socio-ecological model to examine barriers and drivers of demand for childhood vaccination among caregivers in Nigeria, Uganda, and Guinea. It found that the decision to vaccinate a child is informed by a caregiver’s web of family and community relationships and a range of environmental and contextual factors. Many of the findings were consistent with existing knowledge on this topic: the influence of the caregiver’s husband in vaccination decisions, the importance of community norms and the damaging impact of poor healthcare system experiences were reaffirmed (Brown et al., 2010; Cobos Muñoz et al., 2015; Dubé et al., 2013; Falagas & Zarkadoulia, 2008; Favin et al., 2012; Feletto et al., 2018; Kestenbaum & Feemster, 2015; MacDonald, 2015; Mohanty et al., 2018). The study adds to our understanding of vaccine demand by examining the role of the caregiver’s interacting microsystems in detail, rethinking the roles of traditional medicine and religious belief, and questioning the assumption that the decision is primarily in the hands of the child’s mother.

The idea that a caregiver’s level of demand for vaccination results, in part, from the interaction of various microsystems within the mesosystem layer of the Socio-Ecological Model has not previously been described in detail to the best of our knowledge. This study adds the perspective that demand for vaccination is informed by tensions and negotiations within a caregiver’s interpersonal relationships; this may include conflict between their husband and mother-in-law, or a marital dispute that is mediated by community elders. The involvement of several individuals in vaccination decision-making suggests that ‘whole family’ or ‘whole community’ intervention approaches, which encourage entire families or communities to work towards a desired endpoint and have shown some success in other policy areas and settings, could be appropriate to encourage vaccination in Sub-Saharan Africa (Mummery & Brown, 2009; Stanley & Humphreys, 2017). Programmes which employ community elders and leaders to advocate for vaccination are relatively common in this setting, but have met with limited success due to a lack of ongoing training to sustain motivation and engagement (Oku et al., 2017; Oyo-Ita et al., 2021; Warigon et al., 2016). The results of this study contain detailed descriptions of how community leaders inform decision making within families, and so could be used to design more robust community-based programmes.

Previous research has suggested that religious convictions and use of traditional medicine constitute threats to uptake of vaccination (Fourn et al., 2009; Ruijs et al., 2012; Streefland, 2001). According to our study, belief in God’s protection may in some cases lead to a rejection of vaccination, but religious teachings may uphold gender norms that reduce a caregiver’s capacity to demand vaccination in more circuitous ways. Similarly, traditional protection methods can be a direct replacement for vaccination, but using traditional approaches also has an important function in promoting community belonging and cohesion that goes beyond child protection. This means that interventions should work with a community’s religious and traditional norms rather than attempt to circumnavigate or supplant them. Programmes could even attempt to partner with traditional medicine practitioners to encourage vaccination, as has been done in South Africa to encourage adherence to HIV medication (*The AMREF 2012 Annual Report*, 2012).

The study questions two assumptions that are prevalent in the literature on vaccine demand. Firstly, existing frameworks tend to conceptualise the issue as an individual decision on the part of the primary caregiver (Habersaat & Jackson, 2020). We provide evidence that even if the caregiver has responsibility for childhood vaccination, their decision-making is informed by their interpersonal relationships, interactions with social systems and belief in prevailing norms and values. Nevertheless, the work perpetuates this assumption by looking at the issue primarily from the caregiver’s point of view, leaving our theoretical approach open to refinement. Future work could build on it by using models and theories which take a wider and less person-centric approach to further interrogate the role of social and structural forces in creating or undermining vaccination demand.

Secondly, when contextual and environmental factors are examined, they are often considered in isolation rather than in conversation with each other. Using a Socio-Ecological Model suggests that the decision to vaccinate a child can be viewed as the product of a complex relationship between interpersonal, community, institutional and environmental factors which are inextricably linked. This is demonstrated, for example, through the finding that religious beliefs within the macrosystem inform familial relations within the microsystem, a linkage which would not be possible if each domain were considered separately. This understanding can be used to inform the design of more holistic interventions to encourage vaccination uptake, in line with implementation science theories which suggest that interventions have to act on multiple levels to be effective (Stevens et al., 2017).

Further research is required to understand the relative importance of each domain of the Socio-Ecological Model for vaccination demand. This could be achieved through path analysis or structural equation modelling on quantitative datasets.

## Limitations

The study has several limitations which should be considered when interpreting the results. All answers are self-reported and unverified, and so may be affected by social desirability or recall biases. The results may not be generalizable to other settings as the sampling method was designed to ensure we included a cross-section of experiences, rather than to be representative of each country. The incentive offered in Guinea to participants was high (to overcome recruitment challenges), which could have introduced selection bias.

Although we used an established SEM as the basis of the coding, we made some adaptations to best suit our purposes, which have not been validated. The model also does not cover every factor which could impact vaccination demand. For example, it does not include personal experiences of vaccination and vaccine-preventable diseases, which may inform a caregiver’s decision about immunization. This will result in an incomplete view of the determinants of vaccination demand, and so the research should be interpreted in conjunction with other sources.

The analysis process followed a pre-determined protocol, but researcher bias may have affected which subcodes were created and which were reported in this article. None of the coders were from the countries included in the study. Although the results were discussed and interpreted with co-authors from Guinea, Uganda, and Nigeria, this could have introduced western bias into the interpretation of the data.

## Conclusion

Much scholarship on demand for vaccination focuses on primary caregivers’ decision-making processes and does not sufficiently integrate this with contextual and interpersonal forces that may shape their demand for infant immunization. This article concludes that several factors come together to inform a caregiver’s demand for vaccination, including their familial and social relationships, their interactions with government and healthcare institutions, and the wider social and cultural environment in which they live. The work has implications for intervention design and suggests that more holistic approaches could be beneficial in creating and sustaining demand for vaccinations.

## Data Availability

Reasonable data requests will be considered by the authors

## Acknowledgments

We would like to acknowledge the contributions of Holly Exton-Smith, Marcos Fernandez, Virginia Nkwanzi, Alhassane Baldé, Mohamed Dioubaté, Idalecio Agostinho das Neves, Eleanor Tait, Sophie Mathison, Possy Mugyenyi, Lisa Oot, Kate Bagshaw, Rebecca Fields, Ugochukwu Osigwe, Ndadilnasiya Endie Waziri, Yusuf Yusufari, Jenny Sequeira, Sarah Chesemore, Wenfeng Gong, Anna Rapp, Tracy Johnson and Andrew Buhayar during the fieldwork and analysis phases of the study, as well as the moderators and participants involved in the research.

## References

Annual Progress Report (2019). (2019). Gavi, The Vaccine Alliance. https://www.gavi.org/sites/default/files/programmes-impact/our-impact/apr/Gavi-Progress-Report-2019_1.pdf

Bloom, D. E. (2011). The value of vaccination. Advances in Experimental Medicine and Biology, 697, 1–8. https://doi.org/10.1007/978-1-4419-7185-2_1

Bronfenbrenner, U. (1989). Ecological systems theory (Vol. 6). Greenwich, CT: JAI.

Brown, K. F., Kroll, J. S., Hudson, M. J., Ramsay, M., Green, J., Long, S. J., Vincent, C. A., Fraser, G., & Sevdalis, N. (2010). Factors underlying parental decisions about combination childhood vaccinations including MMR: A systematic review. Vaccine, 28(26), 4235–4248. https://doi.org/10.1016/j.vaccine.2010.04.052

Cobos Muñoz, D., Monzón Llamas, L., & Bosch-Capblanch, X. (2015). Exposing concerns about vaccination in low- and middle-income countries: A systematic review. International Journal of Public Health, 60(7), 767–780. https://doi.org/10.1007/s00038-015-0715-6

Dubé, E., Laberge, C., Guay, M., Bramadat, P., Roy, R., & Bettinger, J. A. (2013). Vaccine hesitancy. Human Vaccines & Immunotherapeutics, 9(8), 1763–1773. https://doi.org/10.4161/hv.24657

Falagas, M. E., & Zarkadoulia, E. (2008). Factors associated with suboptimal compliance to vaccinations in children in developed countries: A systematic review. Current Medical Research and Opinion, 24(6), 1719–1741. https://doi.org/10.1185/03007990802085692

Favin, M., Steinglass, R., Fields, R., Banerjee, K., & Sawhney, M. (2012). Why children are not vaccinated: A review of the grey literature. International Health, 4(4), 229–238. https://doi.org/10.1016/j.inhe.2012.07.004

Feikin, D. R., Flannery, B., Hamel, M. J., Stack, M., & Hansen, P. M. (2016). Vaccines for Children in Low- and Middle-Income Countries. In R. E. Black, R. Laxminarayan, M. Temmerman, & N. Walker (Eds.), Reproductive, Maternal, Newborn, and Child Health: Disease Control Priorities, Third Edition (Volume 2). The International Bank for Reconstruction and Development / The World Bank. http://www.ncbi.nlm.nih.gov/books/NBK361927/

Feletto, M., Sharkey, A., Rowley, E., Gurley, N., & Sinha, A. (2018). A Gender Lens to Advance Equity in Immunization (No. 05; ERG Discussion Papers). Equity Reference Group for Immunization.

Fourn, L., Haddad, S., Fournier, P., & Gansey, R. (2009). Determinants of parents’ reticence toward vaccination in urban areas in Benin (West Africa). BMC International Health and Human Rights, 9(Suppl 1), S14. https://doi.org/10.1186/1472-698X-9-S1-S14

Golden, S. D., & Earp, J. A. L. (2012). Social Ecological Approaches to Individuals and Their Contexts: Twenty Years of Health Education & Behavior Health Promotion Interventions. Health Education & Behavior, 39(3), 364–372. https://doi.org/10.1177/1090198111418634

Habersaat, K. B., & Jackson, C. (2020). Understanding vaccine acceptance and demand— And ways to increase them. Bundesgesundheitsblatt - Gesundheitsforschung - Gesundheitsschutz, 63(1), 32–39. https://doi.org/10.1007/s00103-019-03063-0

Hickler, B., MacDonald, N. E., Senouci, K., & Schuh, H. B. (2017). Efforts to monitor Global progress on individual and community demand for immunization: Development of definitions and indicators for the Global Vaccine Action Plan Strategic Objective 2. Vaccine, 35(28), 3515–3519. https://doi.org/10.1016/j.vaccine.2017.04.056

Institut National de la Statistique & ICF. (2019). Guinea Demographic and Health Survey (EDS V) 2016-18. INS/Guinea and ICF. http://dhsprogram.com/pubs/pdf/FR353/FR353.pdf

Jill F. Kilanowski PhD, F., RN, APRN, CPNP. (2017). Breadth of the Socio-Ecological Model. Journal of Agromedicine, 22(4), 295–297. https://doi.org/10.1080/1059924X.2017.1358971

Kestenbaum, L. A., & Feemster, K. A. (2015). Identifying and Addressing Vaccine Hesitancy. Pediatric Annals, 44(4), e71–e75. https://doi.org/10.3928/00904481-20150410-07

Lane, S., MacDonald, N. E., Marti, M., & Dumolard, L. (2018). Vaccine hesitancy around the globe: Analysis of three years of WHO/UNICEF Joint Reporting Form data-2015-2017. Vaccine, 36(26), 3861–3867. https://doi.org/10.1016/j.vaccine.2018.03.063

Levine, O., Lemango, E., Bernson, J., Gurley, N., Rowley, E., & McIlvaine, B. (2018). Tackling Inequities in Immunization Outcomes in Remote Rural Contexts (No. 08; ERG Discussion Papers). Equity Reference Group for Immunization.

MacDonald, N. E. (2015). Vaccine hesitancy: Definition, scope and determinants. Vaccine, 33(34), 4161–4164. https://doi.org/10.1016/j.vaccine.2015.04.036

Majid, U., & Ahmad, M. (2020). The Factors That Promote Vaccine Hesitancy, Rejection, or Delay in Parents. Qualitative Health Research, 30(11), 1762–1776. https://doi.org/10.1177/1049732320933863

Mohanty, S., Carroll-Scott, A., Wheeler, M., Davis-Hayes, C., Turchi, R., Feemster, K., Yudell, M., & Buttenheim, A. M. (2018). Vaccine Hesitancy in Pediatric Primary Care Practices. Qualitative Health Research, 28(13), 2071–2080. https://doi.org/10.1177/1049732318782164

Mummery, W. K., & Brown, W. J. (2009). Whole of community physical activity interventions: Easier said than done. British Journal of Sports Medicine, 43(1), 39–43. https://doi.org/10.1136/bjsm.2008.053629

Muzumdar, J. M., & Cline, R. R. (2009). Vaccine supply, demand, and policy: A primer. Journal of the American Pharmacists Association, 49(4), e87–e99. https://doi.org/10.1331/JAPhA.2009.09007

National Population Commission - NPC & ICF. (2019). Nigeria Demographic and Health Survey 2018—Final Report. NPC and ICF. http://dhsprogram.com/pubs/pdf/FR359/FR359.pdf

Newman, B. M., & Newman, P. R. (2020). Chapter 11—Ecological theories. In B. M. Newman & P. R. Newman (Eds.), Theories of Adolescent Development (pp. 313– 335). Academic Press. https://doi.org/10.1016/B978-0-12-815450-2.00011-5

Oku, A., Oyo-Ita, A., Glenton, C., Fretheim, A., Eteng, G., Ames, H., Muloliwa, A., Kaufman, J., Hill, S., Cliff, J., Cartier, Y., Bosch-Capblanch, X., Rada, G., & Lewin, S. (2017). Factors affecting the implementation of childhood vaccination communication strategies in Nigeria: A qualitative study. BMC Public Health, 17(1), 200. https://doi.org/10.1186/s12889-017-4020-6

Okwo-Bele, J.-M., Conner, R., McIlvaine, B., Rowley, E., & Bernson, J. (2018). Tackling Inequities in Immunization Outcomes in Conflict Contexts (No. 06; ERG Discussion Papers). Equity Reference Group for Immunization.

Oyo-Ita, A., Bosch-Capblanch, X., Ross, A., Oku, A., Esu, E., Ameh, S., Oduwole, O., Arikpo, D., & Meremikwu, M. (2021). Effects of engaging communities in decision-making and action through traditional and religious leaders on vaccination coverage in Cross River State, Nigeria: A cluster-randomised control trial. PLOS ONE, 16(4), e0248236. https://doi.org/10.1371/journal.pone.0248236

Ruijs, W. L. M., Hautvast, J. L. A., van Ijzendoorn, G., van Ansem, W. J. C., van der Velden, K., & Hulscher, M. E. J. L. (2012). How orthodox protestant parents decide on the vaccination of their children: A qualitative study. BMC Public Health, 12, 408. https://doi.org/10.1186/1471-2458-12-408

Stanley, N., & Humphreys, C. (2017). Identifying the key components of a ‘whole family’ intervention for families experiencing domestic violence and abuse. Journal of Gender-Based Violence, 1(1), 99–115. https://doi.org/10.1332/239868017X14913081639164

Stevens, J., Pratt, C., Boyington, J., Nelson, C., Truesdale, K. P., Ward, D. S., Lytle, L., Sherwood, N. E., Robinson, T. N., Moore, S., Barkin, S., Cheung, Y. K., & Murray, D. M. (2017). Multilevel Interventions Targeting Obesity: Research Recommendations for Vulnerable Populations. American Journal of Preventive Medicine, 52(1), 115–124. https://doi.org/10.1016/j.amepre.2016.09.011

Streefland, P. H. (2001). Public doubts about vaccination safety and resistance against vaccination. Health Policy (Amsterdam, Netherlands), 55(3), 159–172. https://doi.org/10.1016/s0168-8510(00)00132-9

Suk, J. E., Paez Jimenez, A., Kourouma, M., Derrough, T., Baldé, M., Honomou, P., Kolie, N., Mamadi, O., Tamba, K., Lamah, K., Loua, A., Mollet, T., Lamah, M., Camara, A. N., & Prikazsky, V. (2016). Post-Ebola Measles Outbreak in Lola, Guinea, January-June 2015(1). Emerging Infectious Diseases, 22(6), 1106–1108. https://doi.org/10.3201/eid2206.151652

The AMREF 2012 Annual Report. (2012). AMREF. https://amref.org/wp-content/uploads/2017/05/Annual-Report-2012.pdf

The Social-Ecological Model: A Framework for Prevention |Violence Prevention|Injury Center|CDC. (2022, January 18). https://www.cdc.gov/violenceprevention/about/social-ecologicalmodel.html

Uganda Bureau of Statistics - UBOS & ICF. (2018). Uganda Demographic and Health Survey 2016. UBOS and ICF. http://dhsprogram.com/pubs/pdf/FR333/FR333.pdf

Warigon, C., Mkanda, P., Muhammed, A., Etsano, A., Korir, C., Bawa, S., Gali, E., Nsubuga, P., Erbeto, T. B., Gerlong, G., Banda, R., Yehualashet, Y. G., & Vaz, R. G. (2016). Demand Creation for Polio Vaccine in Persistently Poor-Performing Communities of Northern Nigeria: 2013-2014. The Journal of Infectious Diseases, 213 Suppl 3, S79–85. https://doi.org/10.1093/infdis/jiv511

Yahya, M. (2007). Polio Vaccines: ‘No Thank You!’ Barriers to Polio Eradication in Northern Nigeria. African Affairs, 106(423), 185–204.

