## Supplement 1 for "Applying a Socio-Ecological Model to Understand Factors Impacting Demand for Childhood Vaccinations in Nigeria, Uganda, and Guinea"

### Applying a Socio-Ecological Model to Understand Factors Impacting Demand for Childhood Vaccinations in Nigeria, Uganda, and Guinea
### Supplementary Materials
### Research Materials

### Nigeria

#### Screening Questionnaire

**BMGF VACCINE DEMAND GENERATION**

**CAREGIVER SCREENER**

Nigeria

Ipsos Healthcare
Ipsos Healthcare, 3 Thomas More Square, London, E1W 1YW, UK
Internal client use only
V6

**Sample structure:**

| | Nigeria |
|---|---|
| Urban | 24 |
| Rural and remote rural | 24 |
| **Total** | **48** |
| **Overlap with Ethnography** | 12 |

**Quotas for regions**

| Nigeria | Sokoto | Lagos | Ebonyi |
|---|---|---|---|
| n= | 16 | 16 | 16 |

**Quotas for vaccination status:**

| | IDIs (spread as evenly as possible across states) | Ethno (spread evenly across states) |
|---|---|---|
| Complete immunization | 12 | 4 (2 Lagos, 2 Sokoto) |
| Partial immunization** | 12 | 4 (2 Lagos, 2 Sokoto) |
| Non-immunisation* | 12 | 4 (2 Lagos, 2 Sokoto) |
| **Total** | **36** | **12** |

**Note:** The exact sample splits may change based on what is feasible or advisable in each country

**Quotas for income level:**

| | IDIs | Ethno |
|---|---|---|
| High income | No more than 6 | No more than 2 |
| Low income | No less than 18 | |

**Quotas for setting:**

| | IDIs | Ethno |
|---|---|---|
| Urban | 18 | 6 |
| Rural | 12 | 4 |
| Remote rural | 6 | 2 |

**Quotas for education level (IDIs):**

| | Lagos | Sokoto | Enugu |
|---|---|---|---|
| No education | | 6 | |
| Primary completed | 3<br>(no OR primary) | 3 | 3<br>(no OR primary) |
| Secondary completed | 6 | 2 | 7 |
| Higher than secondary | 3 | 1 | 2 |
| **TOTAL** | **12** | **12** | **12** |

| Recruitment Script |
|---|
| My name is <name> working on behalf of Ipsos Healthcare. I'd like to see if you would be interested in taking part in a research study on child health.<br><br>This research is being conducted on behalf of the Bill and Melinda Gates Foundation, an international NGO, and has been approved by [xxx]. I would like to ask you some questions about yourself to check whether you qualify. This will only take a few minutes of your time.<br><br>Before I start, I want to assure you that we will not share the information you tell us with anyone else. The aim of this research is to understand what you think. We are not trying to sell you anything. Any information you tell us will be treated in confidence and the answers will not be linked to your name.<br><br>This interview itself will last approximately **90 minutes** and it will be carried out in person, at a time that is convenient for you. If you are one of the types of people we need for this research and you choose to participate, then we would like to offer you [5,000 Naira] as a token of appreciation for your time and contribution. |
| **Privacy** |
| The interview will be audio recorded to help us with our analysis.<br><br>Would you be interested in taking part on this basis? |

| | |
|---|---|
| Yes – Continue | No – Thank and Close |

| | | |
|---|---|---|
| QS1. | **ASK ALL**<br><br>Which languages would you prefer to complete an interview in?<br><br>English<br><br>Igbo<br><br>Yoruba<br><br>Hausa<br><br>Pidgin<br><br>Other (record ___________) | <br><br><br><br>Continue<br><br>Continue<br><br>Continue<br><br>Continue<br><br>Continue<br><br>Continue, but check with Ipsos |
| QS2. | **ASK ALL**<br><br>Do you have any children?<br><br>Yes<br><br>No | <br><br><br><br>Continue<br><br>Close |
| QS3. | **ASK ALL**<br><br>Are you responsible for the day to day care of your children?<br><br>Yes<br><br>No | <br><br><br><br>Continue<br><br>Close |
| QS4. | **ASK ALL**<br><br>How old are each of your children?<br><br>1. _______years OR _______months<br><br>2. _______years OR _______months<br><br>3. _______years OR _______months<br><br>4. _______years OR _______months<br><br>5. _______years OR _______months<br><br>6. _______years OR _______months | <br><br><br><br>**CLOSE IF 0 CHILDREN AGED BETWEEN 2 – 5 YEARS** |

| | | |
|---|---|---|
| | 7. ______years OR _____months<br><br>8. ______years OR ______months<br><br>MODERATOR IDENTIFY THE YOUNGEST CHILD IN THE 2-4 YEAR CATEGORY. SAY: **FOR THE REST OF THE QUESTIONS PLEASE THINK ABOUT YOUR YOUNGEST CHILD WHO IS AGED 2-4** | |
| QS5. | **ASK ALL**<br><br>**I'm going to be asking you questions about vaccines** [*Abere Ajesara* in Yoruba**,** *Ogwu Okun* in Igbo, *Alluran Rigakafi* in Hausa]**. By this I mean something that is given to people, when they are not ill, to strengthen their body's ability to fight certain diseases. Sometimes they are given a vaccine as an injection, but vaccines can also be given by mouth**<br><br>Thinking about this child, did they receive any vaccines before the age of 2? **(use alternative wording as needed)?** | **Check quota, classify 'no' as non-immunisation** |
| | Yes | Continue |
| | No | Continue |
| | Don't know | Continue |
| QS6. | **ASK ALL WHOSE CHILD HAS RECEIVED A VACCINATION OR DON'T KNOW WHETHER THEIR CHILD HAS RECEIVED A VACCINATION AT QS5**<br><br>Do you have a vaccination card for this child? | |
| | Yes | Continue |
| | No | Close IF ANSWERED DON'T KNOW AT QS5 |
| QS6b. | **CHECK VACCINATION CARD OF ALL WHO ANSWERED DON'T KNOW AT S5 AND YES AT S6** | |
| | ALL VACCINATIONS SHOWN<br><br>SOME VACCINATIONS SHOWN<br><br>NO VACCINATIONS SHOWN | ALL - Go to **S10** to record vaccinations and follow instructions on vaccination status allocation |
| QS7. | **ASK ALL WHOSE CHILD HAS RECEIVED A VACCINATION**<br><br>Please think about the times when this child had vaccines. At any of these times… | **Record and classify** |
| | …were there posters about the vaccine around your town? | YES: Campaign |

| | | |
|---|---|---|
| | …was the vaccine mentioned on the radio? | YES: Campaign |
| | …were lots of other children in your area getting the same vaccination on the same day? | YES: Campaign |
| | Did you go for the vaccination because the health facility told you it was time to get that vaccination for your child? | YES: Routine |
| | …were there many vaccinators (rather than just one person)? | YES: Campaign |
| | …Did someone come to your area specifically to do the vaccinations? | YES: Outreach/ Campaign |
| | Did the vaccination take place in a clinic? | Routine/ campaign |
| | Did the vaccination take place in a public place (in a tent/ public hall?) | Outreach/ campaign<br><br>**Must select at least one routine/ outreach option to continue (i.e. not just campaign)** |
| QS8. | **ASK ALL WHOSE CHILD HAS RECEIVED A VACCINATION AND RECRUITER DOES NOT HAVE ACCESS TO THEIR VACCINATION CARD**<br><br>As far as you remember, how many times did you take your child for a vaccination when they were between the ages of 0 and 2 (including any vaccinations they received when they were born)?<br><br>Record visits_________________ | **Visits – capture only** |
| QS9. | **ASK ALL WHOSE CHILD HAS RECEIVED A VACCINATION AND RECRUITER DOES NOT HAVE ACCESS TO THEIR VACCINATION CARD**<br><br>And how many individual vaccines did they receive altogether? This could either be an injection or something they swallow.<br><br>Record number of vaccines_________________ | |
| QS10. | **IF VACCINATION CARD AVALABLE, REFER TO VACCINATION CARD AND RECORD WHICH VACCINATIONS HAVE BEEN ADMINISTERED**<br><br>**Recruiter to complete if access to a vaccination card**<br><br>**Nigeria:** | Record and specify how many doses if several |

| | | |
|---|---|---|
| | Bacille Calmette-Guérin (BCG)<br><br>Hepatitis B paediatric dose<br><br>Oral polio vaccine (OPV)<br><br>Pentavalent (DTwPHibHepB)<br><br>Pneumococcal conjugate vaccine (PCV)<br><br>Inactivated polio vaccine (IPV)<br><br>Measles<br><br>Yellow Fever<br><br>Other | ___________________ *Birth*<br><br>___________________ *Birth*<br><br>___________________ *Birth, 6, 10 & 14 wks*<br><br>___________________ *6, 10 & 14 wks*<br><br>___________________ *6, 10 & 14 wks*<br><br>___________________ *14 wks*<br><br>___________________ *9*<br><br>___________________ *9 months*<br><br>______________________<br>______________________<br>______________________ |
| QS11. | **VACCINATION STATUS**<br><br>Respondents should be categorized into one of these categories by the recruiter:<br><br>**<u>Not vaccinated</u>**<br><br>QS8 – 0 times<br>OR<br>QS10 – nothing on the vaccination card<br><br>**<u>Partially vaccinated</u>**<br><br>QS8 – 1-4 visits<br>OR<br>QS10 – card partially completed<br><br>**<u>Fully vaccinated</u>**<br><br>QS8 – 5 visits<br>OR<br>QS10 – card fully completed | Refer to quotas |

| | | |
|---|---|---|
| QS11b | **ASK IF VACCINATED**<br><br>Did you take your child to private clinics, public clinics, or a mixture of both to get vaccinated?<br><br>All vaccines in private clinic<br><br>Some in private clinic and some in public clinic<br><br>All vaccines in public clinic | <br><br><br><br>Close<br><br>Continue<br><br>Continue |
| QS12. | **ASK ALL**<br><br>How old are you?<br><br>Record age:___________________ | <br><br><br><br>Record |
| QS13. | **RECRUITER TO FILL IN**<br><br>Region<br>Lagos<br><br>Ebonyi<br><br>Sokoto<br><br>Other | <br><br><br>Continue<br><br>Continue<br><br>Continue<br><br>Close |
| QS14. | **RECRUITER TO FILL IN**<br><br>Village/town/city/district<br><br>Record answer: _________________<br><br>Recruiter, Classify:<br><br>Urban<br><br>Rural<br><br>Remote Rural | **Record, classify and check quotas** |
| QS15. | **ASK ALL**<br><br>What is your religion?<br><br>Christian<br>Muslim | **Capture only** |

| | | |
|---|---|---|
| | Traditional spirituality/ beliefs<br>Other religion (please specify) ____________________ | |
| QS16. | **ASK ALL**<br><br>Are you currently working?<br><br>Yes<br><br>No | **Capture only** |
| QS17. | **ASK ALL**<br><br>What is the typical monthly income for your household? | |
| | Over 11.5 million naira | **High- check quotas** |
| | 3.5 to 11.5 million naira | **High- check quotas** |
| | 900,000 to 3.4 million naira | **Middle- check quotas** |
| | 234,000 to 899,999 naira | **Middle- check quotas** |
| | 25,000 to 233,999 naira | **Low- check quotas** |
| | Below 24,999 naira | **Low- check quotas** |
| QS18. | **ASK ALL**<br><br>What is the highest level of education you have completed?<br><br>No education<br>Primary completed<br>Secondary completed<br>Technical College completed<br>University (Bachelors) degree completed<br>Master's degree completed<br>PhD | **CHECK QUOTAS** |
| QS19. | **ASK ALL**<br><br>Finally, we are also looking to speak to a small number of fathers of children, separately to the mothers. Interviews with fathers will last 60 minutes and they will also be offered a token of appreciation for speaking with us.<br><br>Would you be willing to pass on our details to the father of your youngest child and he can contact us to arrange an interview? | |

| | | | |
|---|---|---|---|
| | | Yes<br>No | Continue<br>Continue |

#### Discussion Guide

### VACCINE DEMAND - CAREGIVER DISCUSSION GUIDE:
**90 minutes**

Version 4

INTERNAL / CLIENT USE ONLY

Ipsos Healthcare
3 Thomas More Square, London, E1W 1YW, UK

**Research objectives:**

- Understand the context of the caregiver's life and priorities for them/ their children
- Explore level and nature of influence on decision-making, in general and in relation to their child's vaccination
- Identify beliefs and attitudes towards vaccination
- Map out the caregiver journey on both a functional/chronological level and an emotional one
- Identify leakage points/barriers along this journey
- Identify distinct pathways followed and/or caregiver typologies

Please note:

- **All instructions to the moderator are in bold**

**Materials for the interview (highlighted in yellow throughout):**

**Scenario/ Decision-making exercise cards**
**Definition of vaccination**
**Journey timeline**

#### A. Introduction (2 mins)

**Moderator: Introduce yourself and Ipsos Healthcare**
My name is ___ and I am here representing Ipsos Healthcare, a company that does research all over the world for different types of organisations. Thank you for agreeing to participate in this research. The reason we're talking to caregivers like you is to understand what caregivers think about **child health**. We'll be talking for around 90 minutes.

I want to let you know that:

- I, or the company I work for, is not connected to the government or any clinics. We are independent.
- This research is being done according to international rules on how to do this kind of survey- this is to make sure that you are protected and information about you will be kept safe;
- You have the right to stop the interview at any time.
- Your responses will be combined with other respondents and presented to the Bill and Melinda Gates Foundation together with the responses of other caregivers that we are speaking to;
- The interview will be audio recorded to help us with our analysis only

Are you happy to participate with the interview on this basis?
Yes – **Continue**
No – **Thank you and close**

#### B. Warm-up and introductions (10 mins – total 12 mins)

**MODERATOR GOAL FOR THIS SECTION:** To get to know the respondent, how they live their lives, who they interact with and some of their values and beliefs

**Moderator say:** I'd like to start our discussion today by asking you to introduce yourself.

**Allow spontaneous response, then the following if not mentioned:**

- First name
- Age
- **NOTE FOR MODERATOR: Ensure the respondent is relaxed. If you need an ice breaker then ask:**
  - Just so I can get to know you a bit, please describe to me something which has happened to you this week that stands out to you

##### Family/Domestic Life

I'd like to find out a little bit more about you and your family.

- Tell me about who you live with…
- What is a typical day like for you?

##### Work Life/ Education

- What is your highest level of education?
- Are you currently working? What do you do for work?

##### Personality

- How would those closest to you describe your personality?
  **PROBE FOR AS MUCH DETAIL AS POSSIBLE**
  - Is this also how you would describe yourself?

##### Influences in society

- Who are the people you respect the most (meaning their opinion is important to you)?
- **IF ONLY MENTIONED FAMILY MEMBERS ASK:** What about outside your family?
  - Why do you respect them so much?
- Tell me about a time within the last year where you needed to ask somebody for advice:
  - What was the situation?
  - Who did you go to for advice? Why did you go to that person?
  - Who else do you trust to give you good advice?

##### Religious Identity and Personality

- To what extent does religion play a role in your life?
  - Describe the ways it plays a role in a typical week

Decision-making card exercise: We're now going to look at some short scenarios/stories about parents making decisions. After I have explained the situation, I'd like you to tell me what you think the parent should do.

- Scenario 1

- Scenario 2
- Scenario 3

**MODERATOR, PROBE AROUND REASONS WHY EACH COURSE OF ACTION IS ADVISED BY THE CAREGIVER**

#### C. Journey of health and protection (36 mins – total 48 mins)

| |
|---|
| **Objective:** To elicit attitudes and approaches to general health, from wellbeing and protection to physical and curative health |

Now we're going to talk generally about a range of topics which can affect families

**Aspirations**

Firstly…

- Tell me about your children
- What are your hopes for each of them in the future?

We will now focus on your youngest child who is between the ages of 2 and 4 to understand your experiences and moments in their development during this time. We will talk about all the people such as you, the child's father, their grandparents etc, and the decisions you made or things that you did to ensure the child was in good health and protected during this period.

We will use a piece of paper to define the key stages of the child's development during this time and what you did to look after the child.

Think about your child in their first 3 months of life, between 3-6 months, 6 months – 1 year, 1 year to 18 months and 18 months to 2 years)

Please tell me about **important moments** which happened to your youngest child during the first 3 months of their life. They may be rites of passage like naming ceremonies or circumcision, they may be key moments that showed your child was developing/growing like when they first began to crawl or walk. Let's mark the important moments on this line.

**REPEAT THE ABOVE FOR THE FOLLOWING TIMEFRAMES:**
**-3-6 MONTHS**
**-6 MONTHS-1 YEAR**
**-1 YEAR TO 18 MONTHS**
**-18 MONTHS-2 YEARS**

(**MAKE SURE THE CONVERSATION IS ABOUT THE CHILD CHOSEN AS THE SUBJECT OF THE INTERVIEW: MUST BE THE YOUNGEST CHILD BETWEEN 2-4 YEARS)**

#### Birth

##### ASK IF NOT MENTIONED EARLIER:

- What is your child's first name?
- Why did you choose this name for them?
- Where were they born?
  **CLARIFY IF NEEDED: E.G. AT HOME? AT A HEALTH FACILITY? ANOTHER PLACE?**
- Were all of your children born there? **[IF NO, PROBE FOR REASONS WHY]**
- Who were you with when the child was born?
- Was your child given anything after he/she was born? Tell me what you remember about that… **(PROBE IF NEEDED FOR EACH THING GIVEN: WHO GAVE IT TO THEM, WHY WAS IT GIVEN? WHAT WAS IT? WHAT WAS IT FOR?)**
  - **[IF NOT MENTIONED]** Was your child given any vaccinations? What do you remember about that?

#### Role of caregivers

- Excluding yourself, who were all the main people who took an active role in looking after the wellbeing of **[INSERT CHILD NAME]** in their first 2 years of life?
  - What role did each person play?

#### Protection

- What do you feel **[INSERT NAME OF YOUNGEST CHILD]** should be protected from, in general?
  **PROBE FOR AS MANY AS POSSIBLE**
- Tell me all the ways in which you protect your child…
  **PROBE FOR AS MANY AS POSSIBLE**

#### Health

- How do you know if someone is healthy?
  **PROBE FOR AS MANY ASPECTS OF GOOD HEALTH AS POSSIBLE**
  - What about a child, what are the signs that a child is healthy?
  - What are your views on the health clinics that you have visited?
  - Has anyone ever come to your house or a local meeting place to talk about health, or give medicine?
    - Tell me about that experience
    - What was good about it? What was bad?

- Tell me all of the things you did at each stage of the child's development to ensure that they would be healthy [**Moderator: PLOT THESE ON THE JOURNEY MAP**]

- Apart from the ones given at birth that you told me about, have your children taken any other medications?

    - **IF YES**: Please tell me why they took each medication
    - Where did you get it from?
    - Was it a traditional/herbal treatment or from a pharmacy/clinic/healthcare worker?

**Disease awareness**

- Which diseases are you aware of which can affect children?
- Do you have any experience with any of these diseases?
- Which diseases are you most worried about? why?
- Are there any ways that you can stop your child from getting these diseases? Please tell me about them

#### D. Attitudes to vaccinations (22 mins – total 70 mins)

**Objective:** To understand respondents' attitudes to vaccination and map out health protection journey

**Moderator say:** I'd now like to speak about vaccination and your experiences with it.

##### Awareness of Vaccines and language

- Tell me all the things that you think of when you hear the word 'vaccination'? **ALLOW SPONTANEOUS MENTIONS AND PROBE 'WHAT ELSE'. MAKE A LIST OF THE MENTIONS. THEN FOR EACH MENTION ASK:**
    - Why did that come to mind?
- Can you explain to me what a vaccine is?
    - **[READ OUT IF THEY DON'T KNOW WHAT A VACCINE IS- USE VACCINE EXPLANATION CARD]** Have you heard about this before?
    - How often do you discuss vaccines with other people?
    - What do people say when talking about vaccines? Is it positive or negative?
        - What do they call them?
    - **IF NON VACCINATOR ASK:** Where do people get vaccines? When did you first hear about vaccines?
    - If you wanted to find out more information about vaccines, where would you go or who would you ask?

##### Belief in Vaccines

- What, if any, benefits can you think of about vaccinations?
    - Where did you hear this?
    - Which of these is most important to you? Why?
- What, if any, negative things about vaccinations come to mind?
    - Where did you hear this?
    - Which of these is most worrying for you? Why?
- **IF NOT ALREADY MENTIONED:** Have you received any vaccinations?
    - What do you remember about that?
- **IF NOT ALREADY MENTIONED CHILD'S VACCINATIONS ASK:** Has [**INSERT CHILD FIRST NAME**] had vaccinations before?

**<u>IF YES</u>: ASK TO SEE THE VACCINATION CARD. IF AVAILABLE ASK…**

- Where do you usually store this card?
- How often do you look at this vaccination card?
  - Have you ever been given more than one card for the same child?
  - Would you like this information in another format? What type?

**<u>MARK ALL VACCINES ON THE JOURNEY MAP</u>**

**<u>IF NO CARD BUT CHILD IS VACCINATED ASK</u>:**

- Were you ever given a card?
  - What happened to the card?
  - How do you know which vaccinations the child has had?

There are different ways that children can get vaccinated. It can be through someone coming to your house, all the children of a certain age going to a particular place for vaccination or going to the clinic on a specific date scheduled beforehand.

- Have you experienced any of these for your child?
- Which of these do you prefer? Why?

Tell me about the first vaccination your child had.

- What was the vaccination for (which disease)?
- **IF NOT ALREADY MARKED FROM VACCINATION CARD ASK:** When did it happen?
- How did you learn about where the vaccination was taking place?

- Where did the vaccination take place?
  - At your house/ at a central place in your area where all parents were asked to bring their children on a particular day/ clinic on a specific date scheduled for your child?
  - Was the vaccine oral or injection?

- What do you remember about the journey to get to that place? What was going through your mind on the way there?
  - What were you feeling as the vaccination was taking place?
  - How did the healthcare worker describe or explain the vaccine?
  - What happened after the vaccine was given?
  - What else did the healthcare worker do to the child or advise to you when the vaccine was given?
  - How long did the whole experience take, from…
    - **CLINIC/IN-COMMUNITY VISIT:** when you left your house to when you came back home/
    - **AT HOME VISIT:** from when the health worker arrived at your home to when they left?

**GO THROUGH EACH VACCINATION PROCESS WITH BELOW QUESTIONS**
**IF POSSIBLE, LOOK AT VACCINATION CARD TO ENSURE ALL VACCINATIONS MENTIONED.**

Now, let's talk about the next time you took your child for a vaccination…

- What do you remember about that day?
- Tell me all the ways it was different from the previous time you took your child for a vaccination [before, during and after the vaccination]
  **PROBE ON SPECIFC DETAILS OF PREVIOUS VACCINATIONS TO CONFIRM WHAT WAS DIFFERENT/ THE SAME**

**ONCE GONE THROUGH EACH OF THE VACCINES ASK:**

- What factors encouraged/helped you to get your child vaccinated on each occasion?

- What factors do you think could stop some people from fully vaccinating their children?
  **MAKE LIST OF ALL FACTORS MENTIONED**

- As far as you know, has your child had all the vaccinations they should have had?

**IF NO:**

- What things meant that the child did not have the vaccinations?
  **NOTE: ADD ALL REASONS MENTIONED TO LIST OF POSSIBLE CHALLENGES**

**IF NO TO CHILD EVER BEING VACCINATED BEFORE:**

- What are all the reasons why your child has not received any vaccinations?
  **NOTE: MAKE NOTE OF ALL REASONS MENTIONED**
- What would you have needed in order to have your child vaccinated?

**TO ALL**

- **IF VACCINE SIDE EFFECTS NOT ALREADY MENTIONED ASK:** Have you ever heard people say, or have you ever seen somebody become unwell after being vaccinated?
  - What did you see or hear?
  - How do you feel about it?

#### E. Exploring Leakage Points (15mins – total 85 mins)

**Objective:** Understand emotional impact of leakage points/ barriers and how to tackle them in an ideal world. Evaluate the importance of each of them and what would change if they were resolved.

**MODERATOR SAY:** Earlier you told me about challenges that could have/ did prevent you from getting your child vaccinated.

I'd like to understand these a little more and how they could be made better.

**Moderator: if respondent has vaccinated their child fully, probe instead on reasons why they *could* have missed vaccinations.**

**MODERATOR REFER TO LISTED CHALLENGES/REASON FOR NOT VACCINATING AND SAY:** Here are the challenges you have mentioned so far **[READ OUT LIST]**.

- Is there anything missing?

**MODERATOR, WRITE DOWN ANY ADDITIONAL ONES MENTIONED AT THIS STAGE.**

**MODERATOR SAY:** Now I would like you to order these challenges: Place the one that would be/ has been the biggest challenge at the top, and the one that would be/has been the smallest challenge at the bottom of the table.

**START WITH THE BIGGEST CHALLENGE:**

- Why was this your biggest challenge to getting your child vaccinated?
  **ENCOURAGE RESPONDENT TO LIST ALL THE REASONS. FOR EACH REASON LISTED, BENEFIT LADDER BELOW**
  - Why is **[reason]** a problem for you?
  - What does/would **[insert answer to previous question]** mean for you?

**MODERATOR, REPEAT FOR THE THREE BIGGEST CHALLENGES**

- Now looking at the biggest challenge, how could this be removed or reduced?
- What would this mean for you? For your family?

#### F. Wrap Up (5 mins – total 90 mins)

| |
|---|
| **Objective:** To gauge expectations of the vaccination uptake in the future |

**MODERATOR SAY:** We have just a few minutes at the end here before we finish the discussion.

I am going to read a few sentences and I want you tell me whether you think each one is true or false:

1. In this country, all vaccinations for children are available free of charge
2. In this country, a parent can be sent to prison for not vaccinating their child
3. Vaccinating your child is the best way to keep them from getting sick from some diseases

- Imagine you were in charge of getting every child in your community fully vaccinated, what would you do?
  - **PROBE:** what would you do differently to what is being done these days?

**MODERATOR SAY:** Thank you for all your participation today.

- Is there anything you would like to add, or sum-up from this discussion today?

**Thank and Close**

### Uganda

#### Screening Questionnaire

**BMGF VACCINE DEMAND GENERATION**
**CAREGIVER SCREENER**
Uganda

Ipsos Healthcare
Ipsos Healthcare, 3 Thomas More Square, London, E1W 1YW, UK
Internal client use only
V2

**Sample structure:**

| | IDI | Ethno | Total |
|---|---|---|---|
| **Total** | **18** | **6** | **24** |

**Quotas for regions**

| Uganda | Kampala | Gulu | Mbarara | Total |
|---|---|---|---|---|
| Total | 8 | 8 | 8 | 24 |
| IDI | 6 | 6 | 6 | 18 |
| Ethno | 2 | 2 | 2 | 6 |

**Quotas for vaccination status:**

| | IDIs (spread as evenly as possible across states) | Ethno |
|---|---|---|
| Complete immunization | 6 | 2 |
| Partial immunization | 6 | 2 |
| Non-immunisation | 6 | 2 |
| **Total** | **18** | **6** |

**Note:** The exact sample splits may change based on what is feasible or advisable in each country

**Quotas for income level:**

| | IDIs | Ethno |
|---|---|---|
| High income | No more than 4 | No more than 1 |
| Low income | No less than 9 | No less than 3 |

**Quotas for setting:**

| | IDIs | Ethno |
|---|---|---|
| Urban | 8 | 2 |
| Rural | 6 | 2 |
| Remote rural | 4 | 2 |
| Total | 18 | 6 |

**Quotas for education level (IDIs):**

| Uganda | Kampala | Gulu | Mbarara |
|---|---|---|---|
| No education/primary incomplete | 2 | 2 | 2 |
| Primary completed | 2 | 2 | 2 |
| Secondary completed | 1 | 1 | 1 |
| Higher than secondary | 1 | 1 | 1 |
| **TOTAL** | **6** | **6** | **6** |

**Quotas for education levels (ethnos)**

| Uganda | Total |
|---|---|
| No education/primary incomplete | 2 |
| Primary completed | 2 |
| Secondary completed | 1 |
| Higher than secondary | 1 |
| **TOTAL** | **6** |

| Recruitment Script |
| --- |
| My name is <name> working on behalf of Ipsos Healthcare. I'd like to see if you would be interested in taking part in a research study on child health.<br><br>This research is being conducted on behalf of the Bill and Melinda Gates Foundation, an international NGO, and has been approved by [xxx]. I would like to ask you some questions about yourself to check whether you qualify. This will only take a few minutes of your time.<br><br>Before I start, I want to assure you that we will not share the information you tell us with anyone else. The aim of this research is to understand what you think. We are not trying to sell you anything. Any information you tell us will be treated in confidence and the answers will not be linked to your name.<br><br>We are conducting 5-7 hour long filmed ethnographic interviews. This ethnographic interview will involve a researcher and camera-person spending time with you as you go about a 'normal' day. As they spend time with you, they will ask you some questions about your life – your home, work and family. It will also be carried out at a time that is convenient for you. If you qualify for this research and you choose to participate, then we would like to offer you [xxx] as a token of appreciation for your time and contribution. |
| **Privacy** |
| The interview will be audio recorded to help us with our analysis.<br><br>Would you be interested in taking part on this basis?<br><br>Yes – Continue No – Thank and Close |

| | | |
|---|---|---|
| QS1. | **ASK ALL**<br><br>Which languages would you prefer to complete an interview in? | |
| | English | Continue |
| | Luganda | Continue |
| | Samia | Continue |
| | Japadhola | Continue |
| | Runyankole | Continue |
| | Acholi | Continue |
| | Other (record __________) | Continue, but check with Ipsos |
| QS2. | **ASK ALL**<br><br>Do you have any children? | |
| | Yes | Continue |
| | No | Close |
| QS3. | **ASK ALL**<br><br>Are you responsible for the day to day care of your children? | |
| | Yes | Continue |
| | No | Close |
| QS4. | **ASK ALL**<br><br>How old are each of your children? | |
| | 1. ______years OR ______months | **CLOSE IF 0 CHILDREN AGED BETWEEN 1 AND 2 YEARS** |
| | 2. ______years OR ______months | |
| | 3. ______years OR ______months | |
| | 4. ______years OR ______months | |
| | 5. ______years OR ______months | |

| | | |
|---|---|---|
| | 6. ______years OR ______months<br><br>7. ______years OR _____months<br><br>8. ______years OR ______months<br><br>MODERATOR IDENTIFY THE YOUNGEST CHILD IN THE 2-4 YEAR CATEGORY. SAY: **FOR THE REST OF THE QUESTIONS PLEASE THINK ABOUT YOUR YOUNGEST CHILD WHO IS AGED 2-3** | |
| QS5. | **ASK ALL**<br><br>**I'm going to be asking you questions about vaccines. By this I mean something that is given to people, when they are not ill, to strengthen their body's ability to fight certain diseases. Sometimes they are given a vaccine as an injection, but vaccines can also be given by mouth**<br><br>Thinking about this child, did they receive any vaccines before the age of 2? **(use alternative wording as needed)?** | **Check quota, classify 'no' as non-immunisation** |
| | Yes | Continue |
| | No | Continue |
| | Don't know | Continue |
| QS6. | **ASK ALL WHOSE CHILD HAS RECEIVED A VACCINATION OR DON'T KNOW WHETHER THEIR CHILD HAS RECEIVED A VACCINATION AT QS5**<br><br>Do you have a vaccination card for this child? | |
| | Yes | Continue |
| | No | Close IF ANSWERED DON'T KNOW AT QS5 |
| QS6b. | **CHECK VACCINATION CARD OF ALL WHO ANSWERED DON'T KNOW AT S5 AND YES AT S6** | |
| | ALL VACCINATIONS SHOWN<br><br>SOME VACCINATIONS SHOWN<br><br>NO VACCINATIONS SHOWN | ALL - Go to **S10** to record vaccinations and follow instructions on vaccination status allocation |
| QS7. | **ASK ALL WHOSE CHILD HAS RECEIVED A VACCINATION**<br><br>Please think about the times when this child had vaccines. At any of these times… | **Record and classify** |

| | | |
|---|---|---|
| | …were there posters about the vaccine around your town?<br><br>…was the vaccine mentioned on the radio?<br><br>…were lots of other children in your area getting the same vaccination on the same day?<br><br>Did you go for the vaccination because the health facility told you it was time to get that vaccination for your child?<br><br>…were there many vaccinators (rather than just one person)?<br><br>…Did someone come to your area specifically to do the vaccinations?<br><br>Did the vaccination take place in a clinic?<br><br>Did the vaccination take place in a public place (in a tent/ public hall?) | YES: Campaign<br><br>YES: Campaign<br><br>YES: Campaign<br><br>YES: Routine<br><br>YES: Campaign<br><br>YES: Outreach/ Campaign<br><br>Routine/ campaign<br><br>Outreach/ campaign<br><br>**Must select at least one routine/ outreach option to continue (i.e. not just campaign)** |
| QS8. | **ASK ALL WHOSE CHILD HAS RECEIVED A VACCINATION AND RECRUITER DOES NOT HAVE ACCESS TO THEIR VACCINATION CARD**<br><br>As far as you remember, how many times did you take your child for a vaccination when they were between the ages of 0 and 2 (including any vaccinations they received when they were born)?<br><br>Record visits_________________ | **Visits – capture only** |
| QS9. | **ASK ALL WHOSE CHILD HAS RECEIVED A VACCINATION AND RECRUITER DOES NOT HAVE ACCESS TO THEIR VACCINATION CARD**<br><br>And how many individual vaccines did they receive altogether? This could either be an injection or something they swallow.<br><br>Record number of vaccines_________________ | |
| QS10. | **IF VACCINATION CARD AVALABLE, REFER TO VACCINATION CARD AND RECORD WHICH VACCINATIONS HAVE BEEN ADMINISTERED** | Record and specify how many doses if several |

| | | |
|---|---|---|
| | **Recruiter to complete if access to a vaccination card**<br><br>**Uganda:**<br><br>BCG<br>DTP-HEP-HIB<br>IPV<br>OPV<br>Measles<br>PCV<br>Other | ____________________<br>____________________<br>____________________<br>____________________<br>____________________<br>____________________<br>____________________<br>____________________ |
| QS11. | **VACCINATION STATUS**<br><br>Respondents should be categorized into one of these categories by the recruiter:<br><br>**<u>Not vaccinated</u>**<br><br>QS8 – 0 times<br>OR<br>QS10 – nothing on the vaccination card<br><br>**<u>Partially vaccinated</u>**<br><br>QS8 – 1-4 visits<br>OR<br>QS10 – card partially completed<br><br>**<u>Fully vaccinated</u>**<br><br>QS8 – 5 visits<br>OR<br>QS10 – card fully completed | Refer to quotas |
| QS11b | **ASK IF VACCINATED**<br><br>Did you take your child to private clinics, public clinics, or a mixture of both to get vaccinated?<br><br>All vaccines in private clinic<br><br>Some in private clinic and some in public clinic<br><br>All vaccines in public clinic | <br><br><br><br><br>Close<br><br>Continue<br><br>Continue |
| QS12. | **ASK ALL**<br><br>How old are you? | |

| | | |
|---|---|---|
| | Record age:____________________ | Record |
| QS13. | **RECRUITER TO FILL IN**<br><br>Region<br>UGANDA:<br>Kampala<br><br>Acholi (Gulu)<br><br>Ankole (Mbarara)<br><br>Other | <br><br><br><br>Continue<br><br>Continue<br><br>Continue<br><br>Close |
| QS14. | **RECRUITER TO FILL IN**<br><br>Village/town/city/district<br><br>Record answer: __________________<br><br>Recruiter, Classify:<br><br>Urban<br><br>Rural<br><br>Remote Rural | **Record, classify and check quotas** |
| QS15. | **ASK ALL**<br><br>What is your religion?<br><br>Christian<br>Muslim<br>Traditional spirituality/ beliefs<br>Other religion (please specify) ____________________ | **Capture only** |
| QS16. | **ASK ALL**<br><br>Are you currently working?<br><br>Yes<br><br>No | **Capture only** |
| QS17. | **ASK ALL** | |

| | | |
|---|---|---|
| | What is the typical monthly income for your household?<br><br>Below 2,000,000 ugx<br>2,000,000 - 2,500,000 ugx<br>2,500,001 - 3,000,000 ugx<br>3,000,001 - 3,500,000 ugx<br>3,500,001 - 4,000,000 ugx<br>4,000,001 - 4,500,000 ugx<br>4,500,001 - 5,000,000 ugx<br>5,000,001 - 5,500,000 ugx<br>5,500,001 - 6,000,000 ugx<br>Over 6,000,000 ugx | <br><br>**Low- check quotas**<br>**Middle- check quotas**<br>**Middle- check quotas**<br>**Middle- check quotas**<br>**Middle- check quotas**<br>**Middle- check quotas**<br>**Middle- check quotas**<br>**High- check quotas**<br>**High- check quotas**<br>**High- check quotas** |
| QS18. | **ASK ALL**<br><br>What is the highest level of education you have completed?<br><br>No education<br>Primary completed<br>Secondary completed<br>Technical College completed<br>University (Bachelors) degree completed<br>Master's degree completed<br>PhD | **CHECK QUOTAS** |
| QS19. | **ASK ALL**<br><br>Finally, this research will involve spending time in your home with your family and asking you questions as you go about your day. Would you be comfortable spending a day (5-7 hours) with a researcher and film-maker talking and going about your day?<br><br>Yes<br><br>No<br><br>**Recruiter to evaluate participants based on suitability for** | <br><br><br><br><br>Continue<br><br>CLOSE |

| | |
|---|---|
| | different interview styles. For ethnographic interviews the participant must be open and willing to invite researchers into their home, they also may be more outgoing or articulate. |

#### Discussion Guide- Staggered version

### VACCINE DEMAND - CAREGIVER DISCUSSION GUIDE: Staggered over 4 interviews

Version 3

INTERNAL / CLIENT USE ONLY

Ipsos Healthcare
3 Thomas More Square, London, E1W 1YW, UK

**Research objectives:**

- Understand the context of the caregiver's life and priorities for them/ their children
- Explore level and nature of influence on decision-making, in general and in relation to their child's vaccination
- Identify beliefs and attitudes towards vaccination
- Map out the caregiver journey on both a functional/chronological level and an emotional one
- Identify leakage points/barriers along this journey
- Identify distinct pathways followed and/or caregiver typologies

Please note:

- **All instructions to the moderator are in bold**
- **This master guide contains mini -guides for each of the interview sessions. There are 4 in total and each session is clearly marked with approximate timings for that session**

**Materials for the interview (highlighted in yellow throughout):**

**Scenario/ Decision-making exercise cards**
**Definition of vaccination**
**Journey timeline- use this to help you organise their responses to health activities for the child and vaccines in order**

**BLUE- WEEK 1- Intro and Caregiver context- 20 mins**
**ORANGE- WEEK 2- Journey of health- 25 mins**
**GREEN- WEEK 3- Protection and attitudes to Vaccines- 20 mins**
**PURPLE- WEEK 4- Vaccination Journey and challenges- 30 mins**

### INTERVIEW 1- 20 MINUTES

#### A. Introduction

**Moderator: Introduce yourself and Ipsos Healthcare**
My name is ___ and I am here representing Ipsos Healthcare, a company that does research all over the world for different types of organisations. Thank you for agreeing to participate in this research. The reason we're talking to caregivers like you is to understand what caregivers think about **child health** in order to try to improve experience of health services. We'll be talking for around 90 minutes.

I want to let you know that:

- I, or the company I work for, is not connected to the government or any health centres/ hospitals. We are independent.
- This research has been approved by **the Ministry of Health**
- This research is being done according to international rules on how to do this kind of survey- this is to make sure that you are protected and information about you will be kept safe;
- You have the right to stop the interview at any time.
- Your responses will be combined with other respondents and presented with the responses of other caregivers that we are speaking to;
- The interview will be audio recorded to help us with our analysis only

Are you happy to participate with the interview on this basis?
Yes – **Continue**
No – **Thank you and close**

#### B. Warm-up and introductions

**MODERATOR GOAL FOR THIS SECTION:** To get to know the respondent, how they live their lives, who they interact with and some of their values and beliefs

**Moderator say:** I'd like to start our discussion today by asking you to introduce yourself.

**Allow spontaneous response, then the following if not mentioned:**

- First name
- Age
- **NOTE FOR MODERATOR: Ensure the respondent is relaxed. If you need an ice breaker then ask:**
  - Just so I can get to know you a bit, please describe to me something which has happened to you this week that stands out to you

##### Family/Domestic Life

I'd like to find out a little bit more about you and your family.

- Tell me about who you live with…
- What is a typical day like for you?

##### Work Life/ Education

- What is your highest level of education?
- Are you currently working? What do you do for work?

##### Influences in society

- Tell me about a time within the last year where you needed to ask somebody for advice:
  - What was the situation?
  - Who did you go to for advice? Why did you go to that person?
  - Who else do you trust to give you good advice?

##### Religious Identity

- To what extent does religion play a role in your life?
  - Describe the ways it plays a role in a typical week

##### Decision-making card exercise

We're now going to look at some short scenarios/stories about parents making decisions. After I have explained the situation, I'd like you to tell me what you think the parent should do.

- Scenario 1
- Scenario 2
- Scenario 3

**MODERATOR, PROBE AROUND REASONS WHY EACH COURSE OF ACTION IS ADVISED BY THE CAREGIVER AND WHO ALL THE PEOPLE ARE THAT WOULD BE INVOLVED IN MAKING THE DECISION**

**INTERVIEW 2- 25 MINUTES**

#### C. Journey of health and protection

**Objective:** To elicit attitudes and approaches to general health, from wellbeing and protection to physical and curative health

Now we're going to talk generally about a range of topics which can affect families

##### Aspirations

Firstly…

- Tell me about your children
- What are your hopes for each of them in the future?
  - How will these hopes be achieved?

We will now focus on your youngest child who is between the ages of 1 and 2 to understand your experiences with them.

Let's think specifically about the various things that you or those around you did to ensure that your child would be well and healthy during the first year of their life.

- In the first 3 months of life, what was done to ensure that your child would be well?

**PROBE UNTIL RESPONSES EXHAUSTED. WRITE ALL MENTIONS ON JOURNEY TIMELINE**

**REPEAT THE ABOVE FOR THE FOLLOWING TIMEFRAMES:**
**-3-6 MONTHS**
**-6 MONTHS-1 YEAR**

**(MAKE SURE THE CONVERSATION IS ABOUT THE CHILD CHOSEN AS THE SUBJECT OF THE INTERVIEW: MUST BE THE YOUNGEST CHILD BETWEEN 1-2 YEARS)**

##### Birth

**ASK IF NOT MENTIONED EARLIER:**

- What is your child's first name?
- Why did you choose this name for them?
- Where were they born?
  **CLARIFY IF NEEDED: E.G. AT HOME? AT A HEALTH FACILITY? ANOTHER PLACE?**
- Were all of your children born there? **[IF NO, PROBE FOR REASONS WHY]**
- Who was with you when the child was born?
- Was your child given anything after he/she was born? Tell me what you remember about that… **(PROBE IF NEEDED FOR EACH THING GIVEN: WHO GAVE IT TO THEM, WHY WAS IT GIVEN? WHAT WAS IT? WHAT WAS IT FOR?)**

- Apart from the ones given at birth that you told me about, have your children taken any other medications?
    - **IF YES**: Please tell me why they took each medication
    - Where did you get it from?
    - Was it a traditional/herbal treatment or from a pharmacy/hospital/ health centre/ healthcare worker?

#### Role of caregivers

- Excluding yourself, who were all the main people who took an active role in looking after the wellbeing of **[INSERT CHILD NAME]** in their first year of life?
    - What did each person do?

#### Disease awareness

- Which diseases are you aware of which can affect children?
- Which diseases are you most worried about? why?
- Are there any ways that you can stop your child from getting these diseases? Please tell me about them

#### INTERVIEW 3- 15 MINUTES

##### Protection

- What do you feel **[INSERT NAME OF YOUNGEST CHILD]** should be protected from, in general?
  **PROBE FOR AS MANY AS POSSIBLE**
- Tell me all the ways in which you protect your child…
  **PROBE FOR AS MANY AS POSSIBLE**

##### Health

- What are your views on the health centres/ hospitals/ clinics that you have visited?
- Has anyone ever come to your house or a local meeting place to talk about health, or give medicine?
  - Tell me about that experience
  - What was good about it? What was bad?

#### D. Attitudes to vaccinations

**Objective:** To understand respondents' attitudes to vaccination and map out health protection journey

**Moderator say:** I'd now like to speak about vaccination and your experiences with it.

##### Awareness of Vaccines and language

- Tell me all the things that you think of when you hear the word 'vaccination'?
  **ALLOW SPONTANEOUS MENTIONS AND PROBE 'WHAT ELSE'. MAKE A LIST OF THE MENTIONS. THEN FOR EACH MENTION ASK:**
  - Why did that come to mind?
- Can you explain to me what a vaccine is?
  - **[READ OUT IF THEY DON'T KNOW WHAT A VACCINE IS- USE VACCINE EXPLANATION CARD]** Have you heard about this before?
  - How often do you discuss vaccines with other people?
  - What have you heard people say about vaccines? Is it positive or negative?
    - What do they call them?
  - **IF NON VACCINATOR ASK:** Where do people get vaccines? When did you first hear about vaccines?
  - If you wanted to find out more information about vaccines, where would you go or who would you ask?

##### Belief in Vaccines

- What, if any, good things can you think of about vaccinations?

    - Where did you hear this?
    - Which of these is most important to you? Why?
- What, if any, bad things about vaccinations come to mind?
    - Where did you hear this?
    - Which of these is most worrying for you? Why?
- **IF NOT ALREADY MENTIONED:** Have you received any vaccinations?
    - What do you remember about that?

There are different ways that children can get vaccinated. It can be through someone coming to your house, all the children of a certain age going to a particular place for vaccination or going to the health centre/ hospital on a specific date scheduled beforehand.

- Have you experienced any of these for your child?
- Which of these do you prefer? Why?

#### INTERVIEW 4- 30 MINUTES

We're going to carry on from last week and talk a bit more about vaccinations

- Has [**INSERT CHILD FIRST NAME**] had vaccinations before?

**IF YES: ASK TO SEE THE VACCINATION CARD. IF AVAILABLE ASK…**

- Where do you usually store this card?
- How often do you look at this vaccination card?

**MARK ALL VACCINES ON THE JOURNEY MAP**

**IF NO CARD BUT CHILD IS VACCINATED ASK:**

- Were you ever given a card?
  - What happened to the card?
  - How do you know which vaccinations the child has had?
- Were your child's vaccinations marked in another document? Which one?

Tell me about the first vaccination your child had.

- Tell me all that you remember about that day…
- What was the vaccination for (which disease)?
- **IF NOT ALREADY MARKED FROM VACCINATION CARD ASK:** When did it happen?
- How did you learn about where the vaccination was taking place?
- Where did the vaccination take place?

- What do you remember about the journey to get to the place of vaccination? What was going through your mind on the way there?
  - What were you feeling as the vaccination was taking place?
  - How did the healthcare worker describe or explain the vaccine?
  - What happened after the vaccine was given?
  - What else did the healthcare worker do to the child or advise to you when the vaccine was given?
  - How long did the whole experience take, from…
    - **HEALTH CENTRE/ HOSPITAL/IN-COMMUNITY VISIT:** when you left your house to when you came back home/
    - **AT HOME VISIT:** from when the health worker arrived at your home to when they left?
- What, if anything, was difficult about that day?
  - What are the different things that could have stopped you from getting your child vaccinated on that day?

**IF CANNOT ANSWER SAY:**

- Some people find it difficult to get their child vaccinated. What things do you think could stop some people from vaccinating their child?

**MAKE LIST OF ALL FACTORS MENTIONED**

- What things encouraged/helped you to get your child vaccinated?

**ENCOURAGE THEM TO NAME AS MANY AS RELEVANT FOR THEM**

**GO THROUGH EACH VACCINATION PROCESS WITH BELOW QUESTIONS**
**IF POSSIBLE, LOOK AT VACCINATION CARD TO ENSURE ALL VACCINATIONS MENTIONED.**

Now, let's talk about the next time you took your child for a vaccination…

- What do you remember about that day?
- Tell me all the ways it was different from the previous time you took your child for a vaccination

**PROBE IF NEEDED:**

- What was the journey like on the way to get the vaccine?
- Did you feel any different to how you felt before the previous vaccine?
- What do you remember about the health centre/hospital/venue on that day?
- What was the time with the healthcare worker like?
- What was particularly difficult about that day?
- What helped you complete your child's vaccination on that day?

**ONCE GONE THROUGH VACCINATION PROCESS ASK:**

- As far as you know, has your child had all the vaccinations they should have had?

**IF NO:**

- What things meant that the child did not have the vaccinations?
  **NOTE: ADD ALL REASONS MENTIONED TO LIST OF POSSIBLE CHALLENGES**

**IF NO TO CHILD EVER BEING VACCINATED BEFORE:**

- What are all the reasons why your child has not received any vaccinations?
  **NOTE: MAKE NOTE OF ALL REASONS MENTIONED**
- What would you have needed in order to have your child vaccinated?

#### E. Exploring Leakage Points

**Objective:** Understand emotional impact of leakage points/ barriers and how to tackle them in an ideal world. Evaluate the importance of each of them and what would change if they were resolved.

**MODERATOR SAY:** Earlier you told me about challenges that could have/ did prevent you from getting your child vaccinated.

I'd like to understand these a little more and how they could be made better.

**Moderator: if respondent has vaccinated their child fully, probe instead on reasons why they *could* have missed vaccinations.**

**MODERATOR REFER TO LISTED CHALLENGES/REASON FOR NOT VACCINATING AND SAY:** Here are the challenges you have mentioned so far

**[READ OUT LIST].**

- Is there anything missing?

**MODERATOR, WRITE DOWN ANY ADDITIONAL ONES MENTIONED AT THIS STAGE.**

**MODERATOR SAY:**

- Which one was/would have been your biggest challenge to getting your child vaccinated?

**START WITH THE BIGGEST CHALLENGE:**

- Why was this your biggest challenge to getting your child vaccinated?

  **ENCOURAGE RESPONDENT TO LIST ALL THE REASONS. FOR EACH REASON LISTED, ASK:**
  - Why is **[reason]** a problem for you?
  - What does/would **[insert answer to previous question]** mean for you?

- How could this challenge be removed or reduced?
- What would this mean for you? For your family?

#### F. Wrap Up

**Objective:** To gauge expectations of the vaccination uptake in the future

**MODERATOR SAY:** We have just a few minutes at the end here before we finish the discussion.

I am going to read a few sentences and I want you tell me whether you think each one is true or false:

4. In this country, all vaccinations for children are available free of charge
5. In this country, a parent can be sent to prison for not vaccinating their child
6. Vaccinating your child is the best way to keep them from getting sick from some diseases

- Imagine you were in charge of getting every child in your community fully vaccinated, what would you do?
  - **PROBE:** what would you do differently to what is being done these days?

**MODERATOR SAY:** Thank you for all your participation today.

- Is there anything you would like to add, or sum-up from this discussion today?

**Thank and Close**

#### Discussion guide- Non staggered version

### VACCINE DEMAND - CAREGIVER DISCUSSION GUIDE:
**90 minutes**

Version 3

INTERNAL / CLIENT USE ONLY

Ipsos Healthcare
3 Thomas More Square, London, E1W 1YW, UK

**Research objectives:**

- Understand the context of the caregiver's life and priorities for them/ their children
- Explore level and nature of influence on decision-making, in general and in relation to their child's vaccination
- Identify beliefs and attitudes towards vaccination
- Map out the caregiver journey on both a functional/chronological level and an emotional one
- Identify leakage points/barriers along this journey
- Identify distinct pathways followed and/or caregiver typologies

Please note:

- **All instructions to the moderator are in bold**

**Materials for the interview (highlighted in yellow throughout):**

**Scenario/ Decision-making exercise cards**
**Definition of vaccination**
**Journey timeline- use this to help you organise their responses to health activities for the child and vaccines in order**

#### A. Introduction (2 mins)

**Moderator: Introduce yourself and Ipsos Healthcare**
My name is ___ and I am here representing Ipsos Healthcare, a company that does research all over the world for different types of organisations. Thank you for agreeing to participate in this research. The reason we're talking to caregivers like you is to understand what caregivers think about **child health** in order to try to improve experience of health services. We'll be talking for around 90 minutes.

I want to let you know that:

- I, or the company I work for, is not connected to the government or any health centres/ hospitals. We are independent.
- This research has been approved by **the Ministry of Health**
- This research is being done according to international rules on how to do this kind of survey- this is to make sure that you are protected and information about you will be kept safe;
- You have the right to stop the interview at any time.
- Your responses will be combined with other respondents and presented with the responses of other caregivers that we are speaking to;
- The interview will be audio recorded to help us with our analysis only

Are you happy to participate with the interview on this basis?
Yes – **Continue**
No – **Thank you and close**

#### B. Warm-up and introductions (10 mins – total 12 mins)

| |
|---|
| **MODERATOR GOAL FOR THIS SECTION:** To get to know the respondent, how they live their lives, who they interact with and some of their values and beliefs |

**Moderator say:** I'd like to start our discussion today by asking you to introduce yourself.

**Allow spontaneous response, then the following if not mentioned:**

- First name
- Age
- **NOTE FOR MODERATOR: Ensure the respondent is relaxed. If you need an ice breaker then ask:**
    - Just so I can get to know you a bit, please describe to me something which has happened to you this week that stands out to you

##### Family/Domestic Life

I'd like to find out a little bit more about you and your family.

- Tell me about who you live with…
- What is a typical day like for you?

##### Work Life/ Education

- What is your highest level of education?
- Are you currently working? What do you do for work?

##### Influences in society

- Tell me about a time within the last year where you needed to ask somebody for advice:
    - What was the situation?
    - Who did you go to for advice? Why did you go to that person?
    - Who else do you trust to give you good advice?

##### Religious Identity

- To what extent does religion play a role in your life?
    - Describe the ways it plays a role in a typical week

##### Decision-making card exercise

We're now going to look at some short scenarios/stories about parents making decisions. After I have explained the situation, I'd like you to tell me what you think the parent should do.

- Scenario 1
- Scenario 2
- Scenario 3

**MODERATOR, PROBE AROUND REASONS WHY EACH COURSE OF ACTION IS ADVISED BY THE CAREGIVER AND WHO ALL THE PEOPLE ARE THAT**

**WOULD BE INVOLVED IN MAKING THE DECISION**

#### C. Journey of health and protection (36 mins – total 48 mins)

**Objective:** To elicit attitudes and approaches to general health, from wellbeing and protection to physical and curative health

Now we're going to talk generally about a range of topics which can affect families

**Aspirations**

Firstly…

- Tell me about your children
- What are your hopes for each of them in the future?
  - How will these hopes be achieved?

We will now focus on your youngest child who is between the ages of 1 and 2 to understand your experiences with them.

Let's think specifically about the various things that you or those around you did to ensure that your child would be well and healthy during the first year of their life.

- In the first 3 months of life, what was done to ensure that your child would be well?

**PROBE UNTIL RESPONSES EXHAUSTED. WRITE ALL MENTIONS ON JOURNEY TIMELINE**

**REPEAT THE ABOVE FOR THE FOLLOWING TIMEFRAMES:**
**-3-6 MONTHS**
**-6 MONTHS-1 YEAR**

**(MAKE SURE THE CONVERSATION IS ABOUT THE CHILD CHOSEN AS THE SUBJECT OF THE INTERVIEW: MUST BE THE YOUNGEST CHILD BETWEEN 1-2 YEARS)**

**Birth**

**ASK IF NOT MENTIONED EARLIER:**

- What is your child's first name?
- Why did you choose this name for them?
- Where were they born?
  **CLARIFY IF NEEDED: E.G. AT HOME? AT A HEALTH FACILITY? ANOTHER PLACE?**
- Were all of your children born there? **[IF NO, PROBE FOR REASONS WHY]**
- Who was with you when the child was born?
- Was your child given anything after he/she was born? Tell me what you remember about that… **(PROBE IF NEEDED FOR EACH THING GIVEN: WHO GAVE IT TO THEM, WHY WAS IT GIVEN? WHAT WAS IT? WHAT WAS IT FOR?)**

- Apart from the ones given at birth that you told me about, have your children taken any other medications?
    - **IF YES**: Please tell me why they took each medication
    - Where did you get it from?
    - Was it a traditional/herbal treatment or from a pharmacy/hospital/ health centre/healthcare worker?

#### Role of caregivers

- Excluding yourself, who were all the main people who took an active role in looking after the wellbeing of **[INSERT CHILD NAME]** in their first year of life?
    - What did each person do?

#### Disease awareness

- Which diseases are you aware of which can affect children?
- Which diseases are you most worried about? why?
- Are there any ways that you can stop your child from getting these diseases? Please tell me about them

#### Protection

- What do you feel **[INSERT NAME OF YOUNGEST CHILD]** should be protected from, in general?
  **PROBE FOR AS MANY AS POSSIBLE**
- Tell me all the ways in which you protect your child…
  **PROBE FOR AS MANY AS POSSIBLE**

#### Health

- What are your views on the health centres/ hospitals/ clinics) that you have visited?
- Has anyone ever come to your house or a local meeting place to talk about health, or give medicine?
    - Tell me about that experience
    - What was good about it? What was bad?

#### D. Attitudes to vaccinations (22 mins – total 70 mins)

**Objective:** To understand respondents' attitudes to vaccination and map out health protection journey

**Moderator say:** I'd now like to speak about vaccination and your experiences with it.

#### Awareness of Vaccines and language

- Tell me all the things that you think of when you hear the word 'vaccination'?
  **ALLOW SPONTANEOUS MENTIONS AND PROBE 'WHAT ELSE'. MAKE A LIST OF THE MENTIONS. THEN FOR EACH MENTION ASK:**
    - Why did that come to mind?

- Can you explain to me what a vaccine is?
  **[READ OUT IF THEY DON'T KNOW WHAT A VACCINE IS- USE VACCINE EXPLANATION CARD]**
  Have you heard about this before?
    - How often do you discuss vaccines with other people?
    - What have you heard people say about vaccines? Is it positive or negative?
        - What do they call them?

- **IF NON VACCINATOR ASK:** Where do people get vaccines? When did you first hear about vaccines?
    - If you wanted to find out more information about vaccines, where would you go or who would you ask?

#### Belief in Vaccines

- What, if any, good things can you think of about vaccinations?
    - Where did you hear this?
    - Which of these is most important to you? Why?
- What, if any, bad things about vaccinations come to mind?
    - Where did you hear this?
    - Which of these is most worrying for you? Why?

- **IF NOT ALREADY MENTIONED:** Have you received any vaccinations?
    - What do you remember about that?

- **IF NOT ALREADY MENTIONED CHILD'S VACCINATIONS ASK:**
  Has [**INSERT CHILD FIRST NAME**] had vaccinations before?

**IF YES: ASK TO SEE THE VACCINATION CARD OR FOR THEM TO READ OUT THE VACCINATIONS IN ORDER. IF AVAILABLE ASK… MARK ALL VACCINES ON THE JOURNEY MAP**

- Where do you usually store this card?
- How often do you look at this vaccination card?

**IF NO CARD BUT CHILD IS VACCINATED ASK:**

- Were you ever given a card?
    - What happened to the card?
    - How do you know which vaccinations the child has had?
- Were your child's vaccinations marked in another document? Which one?

There are different ways that children can get vaccinated. It can be through someone coming to your house, all the children of a certain age going to a particular place for vaccination or going to the health centre/hospital on a specific date scheduled beforehand.

- Have you experienced any of these for your child?
- Which of these do you prefer? Why?

Tell me about the first vaccination your child had.

- Tell me all that you remember about that day…
- What was the vaccination for (which disease)?
- **IF NOT ALREADY MARKED FROM VACCINATION CARD ASK:** When did it happen?
- How did you learn about where the vaccination was taking place?
- Where did the vaccination take place?

- What do you remember about the journey to get to the place of vaccination? What was going through your mind on the way there?
    - What were you feeling as the vaccination was taking place?
    - How did the healthcare worker describe or explain the vaccine?
    - What happened after the vaccine was given?
    - What else did the healthcare worker do to the child or advise to you when the vaccine was given?
    - How long did the whole experience take, from…
        - **HEALTH CENTRE/HOSPITAL/IN-COMMUNITY VISIT:** when you left your house to when you came back home/
        - **AT HOME VISIT:** from when the health worker arrived at your home to when they left?
- What, if anything, was difficult about that day?
    - What are the different things that could have stopped you from getting your child vaccinated on that day?

**MAKE A LIST OF FACTORS MENTIONED. IF CANNOT ANSWER SAY:**

- Some people find it difficult to get their child vaccinated. What things do you think could stop some people from vaccinating their child?

**MAKE LIST OF ALL FACTORS MENTIONED**

- What things encouraged/helped you to get your child vaccinated?

**ENCOURAGE THEM TO NAME AS MANY AS RELEVANT FOR THEM**

**GO THROUGH EACH VACCINATION PROCESS WITH BELOW QUESTIONS**
**IF POSSIBLE, LOOK AT VACCINATION CARD TO ENSURE ALL VACCINATIONS MENTIONED.**

Now, let's talk about the next time you took your child for a vaccination…**[REFERENCE THE NEXT TIME THAT THEY MENTIONED E.G. 4 WEEKS]**

- What do you remember about that day?
- Tell me all the ways it was different from the previous time you took your child for a vaccination

**PROBE IF NEEDED:**

- What was the journey like on the way to get the vaccine?
- Did you feel any different to how you felt before the previous vaccine?
- What do you remember about the health centre/ hospital/venue on that day?
- What was the time with the healthcare worker like?
- What was particularly difficult about that day?
- What helped you complete your child's vaccination on that day?

**ONCE GONE THROUGH VACCINATION PROCESS ASK:**

- As far as you know, has your child had all the vaccinations they should have had?

**IF NO:**

- Which ones were missed? (e.g. at what age were they supposed to get it?)
- What things meant that the child did not have the vaccinations?
  **NOTE: ADD ALL REASONS MENTIONED TO LIST OF POSSIBLE CHALLENGES**

**IF NO TO CHILD EVER BEING VACCINATED BEFORE:**

- What are all the reasons why your child has not received any vaccinations?
  **NOTE: MAKE NOTE OF ALL REASONS MENTIONED**
- What would you have needed in order to have your child vaccinated?

#### E. Exploring Leakage Points (15mins – total 85 mins)

**Objective:** Understand emotional impact of leakage points/ barriers and how to tackle them in an ideal world. Evaluate the importance of each of them and what would change if they were resolved.

**MODERATOR SAY:** Earlier you told me about challenges that could have/ did prevent you from getting your child vaccinated.

I'd like to understand these a little more and how they could be made better.

**Moderator: if respondent has vaccinated their child fully, probe instead on reasons why they *could* have missed vaccinations.**

**MODERATOR REFER TO LISTED CHALLENGES/REASON FOR NOT VACCINATING AND SAY:** Here are the challenges you have mentioned so far **[READ OUT LIST]**.

- Is there anything missing?

**MODERATOR, WRITE DOWN ANY ADDITIONAL ONES MENTIONED AT THIS STAGE.**

**MODERATOR SAY:**

- Which one was/would have been your biggest challenge to getting your child vaccinated?

**START WITH THE BIGGEST CHALLENGE:**

- Why was this your biggest challenge to getting your child vaccinated?
  **ENCOURAGE RESPONDENT TO LIST ALL THE REASONS. FOR EACH REASON LISTED, ASK:**
  - Why is **[reason]** a problem for you?
  - What does/would **[insert answer to previous question]** mean for you?

- How could this challenge be removed or reduced?
- What would this mean for you? For your family?

#### F. Wrap Up (5 mins – total 90 mins)

| |
|---|
| **Objective:** To gauge expectations of the vaccination uptake in the future |

**MODERATOR SAY:** We have just a few minutes at the end here before we finish the discussion.

I am going to read a few sentences and I want you tell me whether you think each one is true or false:

7. In this country, all vaccinations for children are available free of charge
8. In this country, a parent can be sent to prison for not vaccinating their child
9. Vaccinating your child is the best way to keep them from getting sick from some diseases

- Imagine you were in charge of getting every child in your community fully vaccinated, what would you do?
  - **PROBE:** what would you do differently to what is being done these days?

**MODERATOR SAY:** Thank you for all your participation today.

- Is there anything you would like to add, or sum-up from this discussion today?

**Thank and Close**

### Guinea

#### Screening Questionnaire

**BMGF VACCINE DEMAND GENERATION**
**CAREGIVER SCREENER**
Guinea

Ipsos Healthcare
Ipsos Healthcare, 3 Thomas More Square, London, E1W 1YW, UK
Internal client use only
V1

**Sample structure:**

| | IDI |
|---|---|
| **Total** | **18** |

**Quotas for regions**

| Guinea | Conakry | Kankan | Mamou | Total |
|---|---|---|---|---|
| IDI | 6 | 6 | 6 | 18 |

**Quotas for vaccination status:**

| | IDIs (spread as evenly as possible across states) |
|---|---|
| Complete immunization | 6 |
| Partial immunization | 6 |
| Non-immunization | 6 |
| **Total** | **18** |

**Quotas for income level:**

| | IDIs |
|---|---|
| High income | No more than 4 |
| Low income | No less than 9 |

**Quotas for setting:**

| | IDIs |
|---|---|
| Urban | 10 |
| Rural | 8 |
| Total | 18 |

**Quotas for education level (IDIs):**

| Guinea | Conakry | Kankan | Mamou |
|---|---|---|---|
| No education/primary incomplete | 2 | 4 | 4 |
| Primary completed | 2 | 1 | 1 |
| Secondary completed | 2 | 1 | 1 |
| Higher than secondary | No more than one across all regions | | |
| **TOTAL** | **6** | **6** | **6** |

| Recruitment Script |
|---|
| My name is <name> working on behalf of Ipsos Healthcare. I'd like to see if you would be interested in taking part in a research study on child health.<br><br>This research is being conducted on behalf of the Bill and Melinda Gates Foundation, an international NGO, and has been approved by [xxx]. I would like to ask you some questions about yourself to check whether you qualify. This will only take a few minutes of your time.<br><br>Before I start, I want to assure you that we will not share the information you tell us with anyone else. The aim of this research is to understand what you think. We are not trying to sell you anything. Any information you tell us will be treated in confidence and the answers will not be linked to your name.<br><br>This interview itself will last approximately **90 minutes** and it will be carried out in person, at a time that is convenient for you. If you are one of the types of people we need for this research and you choose to participate, then we would like to offer you [xxx] as a token of appreciation for your time and contribution. |
| **Privacy** |
| The interview will be audio recorded to help us with our analysis.<br><br>Would you be interested in taking part on this basis?<br><br>Yes – Continue No – Thank and Close |

| | | |
|---|---|---|
| QS1. | **ASK ALL**<br><br>Which languages would you prefer to complete an interview in? | |
| | French | Continue |
| | Soussou | Continue |
| | Malinke | Continue |
| | Peul | Continue |
| | Other (record __________) | Continue, but check with Ipsos |
| QS2. | **ASK ALL**<br><br>Do you have any children? | |
| | Yes | Continue |
| | No | Close |
| QS3. | **ASK ALL**<br><br>Are you responsible for the day to day care of your children? | |
| | Yes | Continue |
| | No | Close |
| QS4. | **ASK ALL**<br><br>How old are each of your children? | |
| | 1. ______years OR ______months | **CLOSE IF 0 CHILDREN AGED BETWEEN 1 AND 3 YEARS** |
| | 2. ______years OR ______months | |
| | 3. ______years OR ______months | |
| | 4. ______years OR ______months | |
| | 5. ______years OR ______months | |
| | 6. ______years OR ______months | |
| | 7. ______years OR _____months | |

| | | |
|---|---|---|
| | 8. ______years OR ______months<br><br>MODERATOR IDENTIFY THE YOUNGEST CHILD IN THE 1-3 YEAR CATEGORY. SAY: **FOR THE REST OF THE QUESTIONS PLEASE THINK ABOUT YOUR YOUNGEST CHILD WHO IS AGED 1-3** | |
| QS5. | **ASK ALL**<br><br>**I'm going to be asking you questions about vaccines. By this I mean something that is given to people, when they are not ill, to strengthen their body's ability to fight certain diseases. Sometimes they are given a vaccine as an injection, but vaccines can also be given by mouth**<br><br>Thinking about this child, did they receive any vaccines before the age of 2? **(use alternative wording as needed)?** | **Check quota, classify 'no' as non-immunisation** |
| | Yes | Continue |
| | No | Continue |
| | Don't know | Continue |
| QS6. | **ASK ALL WHOSE CHILD HAS RECEIVED A VACCINATION OR DON'T KNOW WHETHER THEIR CHILD HAS RECEIVED A VACCINATION AT QS5**<br><br>Do you have a vaccination card for this child? | |
| | Yes | Continue |
| | No | Close IF ANSWERED DON'T KNOW AT QS5 |
| QS6b. | **CHECK VACCINATION CARD OF ALL WHO ANSWERED DON'T KNOW AT S5 AND YES AT S6** | |
| | ALL VACCINATIONS SHOWN<br>SOME VACCINATIONS SHOWN<br>NO VACCINATIONS SHOWN | ALL - Go to **S10** to record vaccinations and follow instructions on vaccination status allocation |
| QS7. | **ASK ALL WHOSE CHILD HAS RECEIVED A VACCINATION**<br><br>Please think about the times when this child had vaccines. At any of these times… | **Record and classify** |
| | …were there posters about the vaccine around your town? | YES: Campaign |
| | …was the vaccine mentioned on the radio? | YES: Campaign |

| | | |
|---|---|---|
| | …were lots of other children in your area getting the same vaccination on the same day?<br><br>Did you go for the vaccination because the health facility told you it was time to get that vaccination for your child?<br><br>…were there many vaccinators (rather than just one person)?<br><br>…Did someone come to your area specifically to do the vaccinations?<br><br>Did the vaccination take place in a clinic?<br><br>Did the vaccination take place in a public place (in a tent/ public hall?) | YES: Campaign<br><br>YES: Routine<br><br>YES: Campaign<br><br>YES: Outreach/ Campaign<br><br>Routine/ campaign<br><br>Outreach/ campaign<br><br>**Must select at least one routine/ outreach option to continue (i.e. not just campaign)** |
| QS8. | **ASK ALL WHOSE CHILD HAS RECEIVED A VACCINATION AND RECRUITER DOES NOT HAVE ACCESS TO THEIR VACCINATION CARD**<br><br>As far as you remember, how many times did you take your child for a vaccination when they were between the ages of 0 and 2 (including any vaccinations they received when they were born)?<br><br>Record visits_________________ | **Visits – capture only** |
| QS9. | **ASK ALL WHOSE CHILD HAS RECEIVED A VACCINATION AND RECRUITER DOES NOT HAVE ACCESS TO THEIR VACCINATION CARD**<br><br>And how many individual vaccines did they receive altogether? This could either be an injection or something they swallow.<br><br>Record number of vaccines_________________ | |
| QS10. | **IF VACCINATION CARD AVALABLE, REFER TO VACCINATION CARD AND RECORD WHICH VACCINATIONS HAVE BEEN ADMINISTERED**<br><br>**Recruiter to complete if access to a vaccination card** | Record and specify how many doses if several |

| | | |
|---|---|---|
| | **Guinea:**<br><br>BCG<br>Oral polio<br>DTC-HEPB-HIB<br>IPV<br>Measles<br>Yellow fever<br>Other | <br><br>____________________<br>____________________<br>____________________<br>____________________<br>____________________<br>____________________<br>____________________<br><br>____________________ |
| QS11. | **VACCINATION STATUS**<br><br>Respondents should be categorized into one of these categories by the recruiter:<br><br>**Not vaccinated**<br><br>QS8 – 0 times<br>OR<br>QS10 – nothing on the vaccination card<br><br>**Partially vaccinated**<br><br>QS8 – 1-4 visits<br>OR<br>QS10 – card partially completed<br><br>**Fully vaccinated**<br><br>QS8 – 5 visits<br>OR<br>QS10 – card fully completed | Refer to quotas |
| QS11b | **ASK IF VACCINATED**<br><br>Did you take your child to private clinics, public clinics, or a mixture of both to get vaccinated?<br><br>All vaccines in private clinic<br><br>Some in private clinic and some in public clinic<br><br>All vaccines in public clinic | <br><br><br><br><br>Close<br><br>Continue<br><br>Continue |
| QS12. | **ASK ALL**<br><br>How old are you?<br><br>Record age:____________________ | <br><br><br><br>Record |

| | | |
|---|---|---|
| QS13. | **RECRUITER TO FILL IN**<br><br>Region<br><br>Conakry<br>Kankan<br>Mamou<br>Other | <br><br><br><br>Continue<br>Continue<br>Continue<br>Close |
| QS14. | **RECRUITER TO FILL IN**<br><br>Village/town/city/district<br><br>Record answer: __________________<br><br>Recruiter, Classify:<br><br>Urban<br>Rural | **Record, classify and check quotas** |
| QS15. | **ASK ALL**<br><br>What is your religion?<br><br>Christian<br>Muslim<br>Traditional spirituality/ beliefs<br>Other religion (please specify) ____________________ | **Capture only** |
| QS16. | **ASK ALL**<br><br>Are you currently working?<br><br>Yes<br>No | **Capture only** |
| QS17. | **ASK ALL**<br><br>What is the typical monthly income for your household?<br><br>0 to 1,967,083 FG<br>1,967,084 to 4,999,999 FG | <br><br><br><br>**Low**<br>**Medium** |

| | | |
|---|---|---|
| | 5,000,000 FG and above | **High** |
| QS18. | **ASK ALL**<br><br>What is the highest level of education you have completed?<br><br>No education<br>Primary completed<br>Secondary completed<br>Technical College completed<br>University (Bachelors) degree completed<br>Master's degree completed<br>PhD | **CHECK QUOTAS** |
| QS19. | **ASK ALL**<br><br>We are also looking to speak to a small number of other family members. Interviews with these people will last 60 minutes and they will also be offered a token of appreciation for speaking with us.<br><br>Would you be willing to pass on our details to others in your family so that we can arrange an interview?<br><br>Yes<br>No | <br><br><br><br><br><br><br>Continue<br>Continue |

#### Discussion Guide

### VACCINE DEMAND - CAREGIVER DISCUSSION GUIDE:
**90 minutes**

Version 1

INTERNAL / CLIENT USE ONLY

Ipsos Healthcare
3 Thomas More Square, London, E1W 1YW, UK

**Research objectives:**

- Understand the context of the caregiver's life and priorities for them/ their children
- Explore level and nature of influence on decision-making, in general and in relation to their child's vaccination
- Identify beliefs and attitudes towards vaccination
- Map out the caregiver journey on both a functional/chronological level and an emotional one
- Identify leakage points/barriers along this journey
- Identify distinct pathways followed and/or caregiver typologies

Please note:

- **All instructions to the moderator are in bold**

**Materials for the interview (highlighted in yellow throughout):**

**Scenario/ Decision-making exercise cards**

**Definition of vaccination**

#### A. Introduction (2 mins)

**Moderator: Introduce yourself and Ipsos Healthcare**
My name is ___ and I am here representing Ipsos Healthcare, a company that does research all over the world for different types of organisations. Thank you for agreeing to participate in this research. The reason we're talking to caregivers like you is to understand what caregivers think about **child health** in order to try to improve experience of health services. We'll be talking for around 90 minutes.

I want to let you know that:

- I, or the company I work for, is not connected to the government or any health centres/ hospitals. We are independent.
- This research has been approved by **the Ministry of Health**
- This research is being done according to international rules on how to do this kind of survey- this is to make sure that you are protected and information about you will be kept safe;
- You have the right to stop the interview at any time.
- Your responses will be combined with other respondents and presented with the responses of other caregivers that we are speaking to;
- The interview will be audio recorded to help us with our analysis only

Are you happy to participate with the interview on this basis?
Yes – **Continue**
No – **Thank you and close**

#### B. Warm-up and introductions (10 mins – total 12 mins)

**MODERATOR GOAL FOR THIS SECTION:** To get to know the respondent, how they live their lives, who they interact with and some of their values and beliefs

**Moderator say:** I'd like to start our discussion today by asking you to introduce yourself.

**Allow spontaneous response, then the following if not mentioned:**

- First name
- Age
- **NOTE FOR MODERATOR: Ensure the respondent is relaxed. If you need an ice breaker then ask:**
    - Just so I can get to know you a bit, please describe to me something which has happened to you this week that stands out to you

##### Family/Domestic Life

I'd like to find out a little bit more about you and your family.

- Tell me about who you live with…
- What role does each member of your family have?
- What is a typical day like for you? [**IF ASK ABOUT COVID PROMPT ON DIFFERENCES BETWEEN PRE/POST]**
- Has this changed since COVID-19?

##### Work Life/ Education

- What is your highest level of education?
- Are you currently working? What do you do for work? [**IF ASK ABOUT COVID PROMPT ON DIFFERENCES BETWEEN PRE/POST]**
- Has this changed since COVID-19?

##### Influences in society

- Tell me about a time within the last year where you needed to ask somebody for advice:
    - What was the situation?
    - Who did you go to for advice? Why did you go to that person?
    - Who else do you trust to give you good advice?

##### Religious Identity

- To what extent does religion play a role in your life?
    - Describe the ways it plays a role in a typical week

##### Decision-making card exercise

We're now going to look at some short scenarios/stories about parents making decisions. After I have explained the situation, I'd like you to tell me what you think the parent should do.

- Scenario 1

- Scenario 2
- Scenario 3

**MODERATOR, PROBE AROUND REASONS WHY EACH COURSE OF ACTION IS ADVISED BY THE CAREGIVER AND WHO ALL THE PEOPLE ARE THAT WOULD BE INVOLVED IN MAKING THE DECISION**

#### C. Journey of health and protection (36 mins – total 48 mins)

**Objective:** To elicit attitudes and approaches to general health, from wellbeing and protection to physical and curative health

Now we're going to talk generally about a range of topics which can affect families

##### Aspirations

Firstly…

- Tell me about your children
- What are your hopes for each of them in the future?
    - How will these hopes be achieved?

We will now focus on your youngest child who is between the ages of 1 and 3 to understand your experiences with them.

##### Birth

**ASK IF NOT MENTIONED EARLIER:**

- What is your child's first name?
- Why did you choose this name for them?
- Where were they born?
  **CLARIFY IF NEEDED: E.G. AT HOME? AT A HEALTH FACILITY? ANOTHER PLACE?**
- Were all of your children born there? **[IF NO, PROBE FOR REASONS WHY]**
- Who was with you when the child was born?
- Was your child given anything after he/she was born? Tell me what you remember about that… **(PROBE IF NEEDED FOR EACH THING GIVEN: WHO GAVE IT TO THEM, WHY WAS IT GIVEN? WHAT WAS IT? WHAT WAS IT FOR?)**

- Apart from the ones given at birth that you told me about, have your children taken any other medications?
    - **IF YES**: Please tell me why they took each medication
    - Where did you get it from?
    - Was it a traditional/herbal treatment or from a pharmacy/hospital/ health centre/healthcare worker/ 'street doctor'?

Let's think specifically about the various things that you or those around you did to ensure that your child would be well and healthy during the first year of their life.

- In the first 3 months of life, what was done to ensure that your child would be well?

**PROBE UNTIL RESPONSES EXHAUSTED. WRITE ALL MENTIONS ON JOURNEY TIMELINE**

**REPEAT THE ABOVE FOR THE FOLLOWING TIMEFRAMES:**
**-3-6 MONTHS**
**-6 MONTHS-1 YEAR**

**(MAKE SURE THE CONVERSATION IS ABOUT THE CHILD CHOSEN AS THE SUBJECT OF THE INTERVIEW: MUST BE THE YOUNGEST CHILD BETWEEN 1-2 YEARS)**

##### Role of caregivers

- Excluding yourself, who were all the main people who took an active role in looking after the wellbeing of **[INSERT CHILD NAME]** in their first year of life?
    - What did each person do?

##### Disease awareness

- Which diseases are you aware of which can affect children?
- Which diseases are you most worried about? why?
- Are there any ways that you can stop your child from getting these diseases? Please tell me about them

##### Protection

- What do you feel **[INSERT NAME OF YOUNGEST CHILD]** should be protected from, in general?
  **PROBE FOR AS MANY AS POSSIBLE**
- Tell me all the ways in which you protect your child…
  **PROBE FOR AS MANY AS POSSIBLE**

##### Health

- Tell us about where you go when you are unwell/ not feeling fine (**PROBE IF NEEDED**: traditional vs health centres/ hospitals)
    - In which circumstances would you go to a traditional healer?
    - And in which circumstances would you go to a health facilities (hospital/ health centre)?
- What are your views on the health centres/ hospitals/ clinics) that you have visited?
- Have you always had that opinion, or has it changed over time?

- Has anyone ever come to your house or a local meeting place to talk about health, or give medicine?
  - Tell me about that experience
  - What was good about it? What was bad?
- To what extent (if at all) do you trust the government to provide health services? Why/ why not?
- To what extent (if at all) would you trust international organisations to provide health services? Why/ why not?

#### D. Attitudes to vaccinations (22 mins – total 70 mins)

**Objective:** To understand respondents' attitudes to vaccination and map out health protection journey

**Moderator say:** I'd now like to speak about vaccination and your experiences with it.

##### Awareness of Vaccines and language

- Tell me all the things that you think of when you hear the word 'vaccination'? **ALLOW SPONTANEOUS MENTIONS AND PROBE 'WHAT ELSE'. MAKE A LIST OF THE MENTIONS. THEN FOR EACH MENTION ASK:**
  - Why did that come to mind?

- Can you explain to me what a vaccine is? **[READ OUT IF THEY DON'T KNOW WHAT A VACCINE IS- USE VACCINE EXPLANATION CARD]** Have you heard about this before?
  - How often do you discuss vaccines with other people?
  - What have you heard people say about vaccines? Is it positive or negative?
    - What do they call them?

- **IF NON VACCINATOR ASK:** Where do people get vaccines? When did you first hear about vaccines?
  - If you wanted to find out more information about vaccines, where would you go or who would you ask?

##### Belief in Vaccines

- What, if any, good things can you think of about vaccinations?
  - Where did you hear this?
  - Which of these is most important to you? Why?
- What, if any, bad things about vaccinations come to mind?
  - Where did you hear this?
  - Which of these is most worrying for you? Why?

- **IF NOT ALREADY MENTIONED:** Have you received any vaccinations?

- What do you remember about that?

- **IF NOT ALREADY MENTIONED CHILD'S VACCINATIONS ASK:**
  Has [**INSERT CHILD FIRST NAME**] had vaccinations before?

**IF YES: ASK TO SEE THE VACCINATION CARD OR FOR THEM TO READ OUT THE VACCINATIONS IN ORDER. IF AVAILABLE ASK…**

- Where do you usually store this card?
- How often do you look at this vaccination card?

**IF NO CARD BUT CHILD IS VACCINATED ASK:**

- Were you ever given a card?
  - What happened to the card?
  - How do you know which vaccinations the child has had?
- Were your child's vaccinations marked in another document? Which one?

There are different ways that children can get vaccinated. It can be through someone coming to your house, all the children of a certain age going to a particular place for vaccination or going to the health centre/hospital on a specific date scheduled beforehand.

- Have you experienced any of these for your child?
- Which of these do you prefer? Why?

Tell me about the first vaccination your child had.

- Tell me all that you remember about that day…
- What was the vaccination for (which disease)?
- **IF NOT ALREADY MARKED FROM VACCINATION CARD ASK:** When did it happen?
- How did you learn about where the vaccination was taking place?
- Where did the vaccination take place?

- What do you remember about the journey to get to the place of vaccination? What was going through your mind on the way there?
  - What were you feeling as the vaccination was taking place?
  - How did the healthcare worker describe or explain the vaccine?
  - What happened after the vaccine was given?
  - What else did the healthcare worker do to the child or advise to you when the vaccine was given?
  - How long did the whole experience take, from…
    - **HEALTH CENTRE/HOSPITAL/IN-COMMUNITY VISIT:** when you left your house to when you came back home/
    - **AT HOME VISIT:** from when the health worker arrived at your home to when they left?
- What, if anything, was difficult about that day?
  - What are the different things that could have stopped you from getting your child vaccinated on that day?

**MAKE A LIST OF FACTORS MENTIONED. IF CANNOT ANSWER SAY:**

- Some people find it difficult to get their child vaccinated. What things do you think could stop some people from vaccinating their child?

**MAKE LIST OF ALL FACTORS MENTIONED**

- What things encouraged/helped you to get your child vaccinated?

**ENCOURAGE THEM TO NAME AS MANY AS RELEVANT FOR THEM**

**GO THROUGH EACH VACCINATION PROCESS WITH BELOW QUESTIONS**
**IF POSSIBLE, LOOK AT VACCINATION CARD TO ENSURE ALL VACCINATIONS MENTIONED.**

Now, let's talk about the next time you took your child for a vaccination…**[REFERENCE THE NEXT TIME THAT THEY MENTIONED E.G. 4 WEEKS]**

- What do you remember about that day?
- Tell me all the ways it was different from the previous time you took your child for a vaccination

**PROBE IF NEEDED:**

- What was the journey like on the way to get the vaccine?
- Did you feel any different to how you felt before the previous vaccine?
- What do you remember about the health centre/ hospital/venue on that day?
- What was the time with the healthcare worker like?
- What was particularly difficult about that day?
- What helped you complete your child's vaccination on that day?

**ONCE GONE THROUGH VACCINATION PROCESS ASK:**

- As far as you know, has your child had all the vaccinations they should have had?

**IF NO:**

- Which ones were missed? (e.g. at what age were they supposed to get it?)
- What things meant that the child did not have the vaccinations?
  **NOTE: ADD ALL REASONS MENTIONED TO LIST OF POSSIBLE CHALLENGES**

**<u>IF NO TO CHILD EVER BEING VACCINATED BEFORE:</u>**

- What are all the reasons why your child has not received any vaccinations?
  **NOTE: MAKE NOTE OF ALL REASONS MENTIONED**
- What would you have needed in order to have your child vaccinated?

#### E. Exploring Leakage Points (15mins – total 85 mins)

**Objective:** Understand emotional impact of leakage points/ barriers and how to tackle them in an ideal world. Evaluate the importance of each of them and what would change if they were resolved.

**MODERATOR SAY:** Earlier you told me about challenges that could have/ did prevent you from getting your child vaccinated.

I'd like to understand these a little more and how they could be made better.

**Moderator: if respondent has vaccinated their child fully, probe instead on reasons why they *could* have missed vaccinations.**

**MODERATOR REFER TO LISTED CHALLENGES/REASON FOR NOT VACCINATING AND SAY:** Here are the challenges you have mentioned so far **[READ OUT LIST]**.

- Is there anything missing?

**MODERATOR, WRITE DOWN ANY ADDITIONAL ONES MENTIONED AT THIS STAGE.**

**MODERATOR SAY:**

- Which one was/would have been your biggest challenge to getting your child vaccinated?

**START WITH THE BIGGEST CHALLENGE:**

- Why was this your biggest challenge to getting your child vaccinated?
  **ENCOURAGE RESPONDENT TO LIST ALL THE REASONS. FOR EACH REASON LISTED, ASK:**
  - Why is **[reason]** a problem for you?
  - What does/would **[insert answer to previous question]** mean for you?

- How could this challenge be removed or reduced?
- What would this mean for you? For your family?

#### F. Wrap Up (5 mins – total 90 mins)

| |
|---|
| **Objective:** To gauge expectations of the vaccination uptake in the future |

**MODERATOR SAY:** We have just a few minutes at the end here before we finish the discussion.

I am going to read a few sentences and I want you tell me whether you think each one is true or false:

10. In this country, all vaccinations for children are available free of charge
11. In this country, a parent can be sent to prison for not vaccinating their child
12. Vaccinating your child is the best way to keep them from getting sick from some diseases

- Imagine you were in charge of getting every child in your community fully vaccinated, what would you do?
    - **PROBE:** what would you do differently to what is being done these days?

**MODERATOR SAY:** Thank you for all your participation today.

- Is there anything you would like to add, or sum-up from this discussion today?

**Thank and Close**
